# Rare penetrant mutations confer severe risk of common diseases

**DOI:** 10.1101/2023.05.01.23289356

**Authors:** Petko Fiziev, Jeremy McRae, Jacob C. Ulirsch, Jacqueline S. Dron, Tobias Hamp, Yanshen Yang, Pierrick Wainschtein, Zijian Ni, Joshua G. Schraiber, Hong Gao, Dylan Cable, Yair Field, Francois Aguet, Marc Fasnacht, Ahmed Metwally, Jeffrey Rogers, Tomas Marques-Bonet, Heidi L. Rehm, Anne O’Donnell-Luria, Amit V. Khera, Kyle Kai-How Farh

## Abstract

We examined 454,712 exomes for genes associated with a wide spectrum of complex traits and common diseases and observed that rare, penetrant mutations in genes implicated by genome-wide association studies confer ∼10-fold larger effects than common variants in the same genes. Consequently, an individual at the phenotypic extreme and at the greatest risk for severe, early-onset disease is better identified by a few rare penetrant variants than by the collective action of many common variants with weak effects. By combining rare variants across phenotype-associated genes into a unified genetic risk model, we demonstrate superior portability across diverse global populations compared to common variant polygenic risk scores, greatly improving the clinical utility of genetic-based risk prediction.

**One sentence summary:** Rare variant polygenic risk scores identify individuals with outlier phenotypes in common human diseases and complex traits.

---

Genome-wide association studies (GWASs) have convincingly identified tens of thousands of common variants underlying complex human traits and diseases (*1*), although several key challenges remain. First, pinpointing which genes these predominately non-coding variants affect is non-trivial, hindering biological insight into disease mechanisms. Second, individual common variants have modest effects on disease risk, resulting in weak aggregate predictors with limited clinical utility and portability between populations (*2–4*). In contrast to GWASs, rare coding variant studies directly link perturbed gene function to specific phenotypes. For individuals with cancer or rare genetic diseases, analysis of whole exome sequencing (WES) routinely uncovers rare, highly penetrant variants that can dramatically alter the course of clinical management (*5–8*) and drive treatment decisions (*9, 10*). However, in the context of common diseases, the role of rare coding variants has not been established to the same extent due to lack of methods for accurately predicting variant function and insufficient cohort sizes.

Recent large-scale genome and exome sequencing studies of the general population have revealed that the average person carries dozens of potentially deleterious rare variants that have arisen through recent germline mutation (*11*). These studies provide the opportunity to move beyond rare genetic disease and examine the impact of medium-to-large effect rare coding variants on a comprehensive set of complex human traits and diseases. In practice, individually rare variants are often combined into burden tests to more powerfully discover genes underlying these phenotypes, but these tests are limited by our ability to distinguish pathogenic from benign variants. Here, we show that our recently developed method PrimateAI-3D (*12*), a 3-D convolutional neural network trained on common genetic variants from 233 primate species, accurately quantifies missense variant pathogenicity, resulting in improved gene discovery across 454,712 individuals in the UK Biobank (*13–15*). We then show how rare variants in these genes can be combined into a unified genetic risk score which has distinct advantages over common variant polygenic risk scores, offering a glimpse into the potential utility of personal genome sequencing for the general population.

## PrimateAI-3D empowers gene discovery in rare variant association tests

To identify genes underlying complex human traits and diseases, we performed rare variant burden tests for 90 well-powered, non-redundant clinical and quantitative phenotypes, including both medical diagnoses and commonly measured laboratory tests, for 454,712 individuals in the UK Biobank who underwent WES (Table S1-S3) (*16*). Using an allele frequency (AF) threshold of 0.1%, we detected 1,841 gene-phenotype associations with loss-of-function (LoF) variants, 1,510 associations with missense variants, and 3,035 associations combining missense and LoF variants (average of 33.7 per phenotype) at a false discovery rate (FDR) of 5% (**Fig. 1A**). When we applied PrimateAI-3D (*12*) to classify pathogenic and benign missense variants, we improved gene discovery by 73%, identifying 1,285 more gene-phenotype associations at the same FDR (**Fig. 1A**, **Fig. S1**, and Table S4). As a negative control, we repeated the test considering rare synonymous variants but detected only 28 gene-phenotype associations. Taken together, these results show that our rare variant tests are well calibrated and that PrimateAI-3D pathogenicity predictions improve gene discovery.

**Fig. 1.**
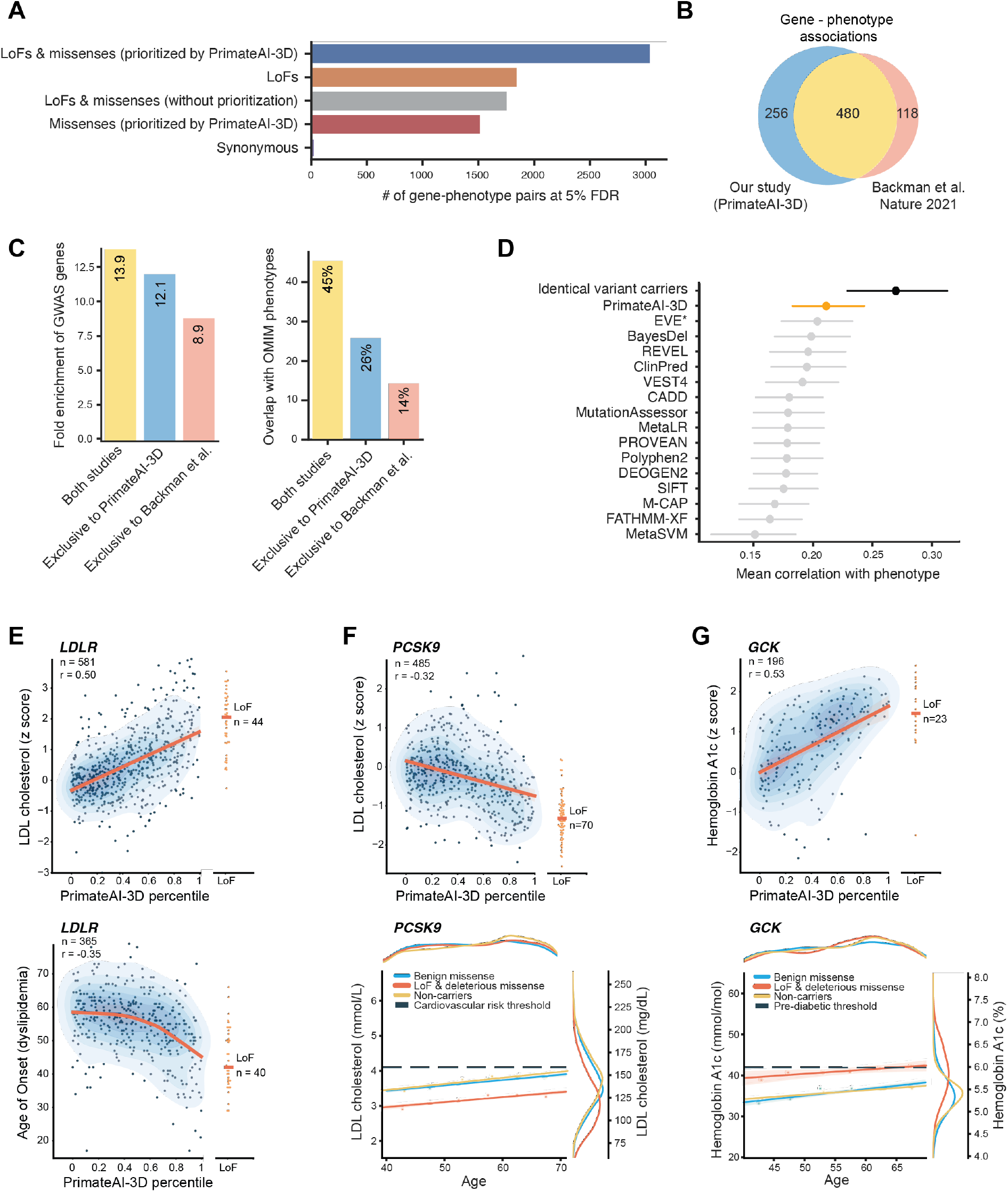
PrimateAI-3D identifies rare deleterious variants that affect disease severity and age of onset. **(A)** Total number of significant gene-phenotype associations (FDR < 5%) identified across 90 phenotypes for rare variant burden tests using different inclusion criteria for variants. As a negative control, the number of significant genotype-phenotype associations for a burden test using only synonymous variants is also shown. (**B**) Comparison of the current study with a recent study of rare variants in the UK Biobank (*17*) on the number of gene-phenotype associations detected exclusively by one or both studies for the same traits and matched significance thresholds. **(C)** Comparison of rare variant genes discovered in this study versus the previous study (*17*) using orthogonal genetic evidence. (**left**) Fold enrichment of rare variant genes at common variant GWAS loci, matched for the same phenotypes. (**right**) Percentage of rare variant genes overlapping with OMIM genes matched for related phenotypes. (**D**) Performance of different variant pathogenicity classifiers (see Methods) at predicting variant effects on quantitative phenotypes. Spearman correlations between pathogenicity scores and phenotype values on a set of 62 gene-phenotype pairs are shown. The phenotypic correlation between individuals carrying an identical missense variant is shown in black as an upper bound for classifier performance. Dots and error bars represent mean ± 95% confidence interval. **(E)** (**top**) Positive correlation of LDL cholesterol concentrations (y-axis) with PrimateAI-3D scores (x-axis) for rare missense variants in *LDLR*. (**bottom)** PrimateAI-3D score is predictive of age of onset for dyslipidemia in carriers of rare missense variants in *LDLR*. (**F**) (**top**) Negative correlation of LDL cholesterol concentrations with PrimateAI-3D scores for rare missense variants in *PCSK9*, a down-regulator of *LDLR*. (**bottom**) LDL cholesterol concentrations increase with age at a similar rate regardless of carrier status, but carriers of prioritized rare variants have lower LDL concentrations across all ages. (**G**) (**top**) Positive correlation of HbA1c concentrations with PrimateAI-3D scores for rare missense variants in *GCK*. (**bottom**) HbA1c concentrations increase with age at a similar rate regardless of carrier status, but carriers of rare deleterious variants reach pre-diabetic thresholds earlier in their lives on average. Deleterious and benign missense variants are defined as variants with PrimateAI-3D score greater than 0.5 and less than 0.5, respectively. For **E**, **F** and **G** red, blue or yellow lines show regression models fitted to the data.

We undertook several additional approaches to validate our gene-phenotype associations and to compare to prior efforts. First, we investigated the strength of support from common variant studies for the gene-phenotype pairs identified by our approach. After performing matched GWASs for the 90 phenotypes (Table S5) (*16*), we observed that 70% of the 3,035 gene-phenotype pairs had a significant GWAS variant (P < 5×10^−8^) within 1 megabase of the transcription start site. Next, we compared our results to a recent rare variant association study in the same UK Biobank cohort (*17*) (**Fig. 1B**). Backman et al. used a burden test which included all LoF variants but permitted only missense variants predicted to be deleterious by five commonly used missense pathogenicity classifiers (*18*). For matched phenotypes and significance thresholds (*16*), we identified 23% more gene-phenotype pairs (Table S6). Gene-phenotype pairs identified exclusively in the present study were more enriched for genes implicated by matching GWASs and overlapped more with genes in related Mendelian diseases (**Fig. 1C**, Table S7), which supports their relevance to complex trait biology. Third, we benchmarked PrimateAI-3D against 15 other pathogenicity classifiers by integrating them into our burden testing pipeline. Again, gene-phenotype pairs detected exclusively by PrimateAI-3D had consistently higher enrichments for GWAS genes for the same trait compared to any other method (**Fig. S2**). Finally, we assessed how well each classifier could predict the effect size of individual variants on phenotype across 62 gene-phenotype pairs detected without variant prioritization (Table S8) (*16*) and again observed that PrimateAI-3D outperformed all other methods (median Wilcoxon P=8×10^−7^, **Fig. 1D, Fig. S3**).

Having comprehensively validated our use of PrimateAI-3D for rare variant burden testing, we explored the correlations we observed between PrimateAI-3D scores, clinical laboratory measurements, and ages of onset for common diseases. In general, we observed a linear relationship with the quantitative measurements and an inverse correlation with age of disease onset (Table S9). We focus on the examples of *LDLR* and *PCSK9* with low-density lipoprotein (LDL) cholesterol levels and *GCK* with glycated hemoglobin A1c (HbA1c) to illustrate these general findings (**Fig 1E-G**). Overall, 1,307 individuals (0.3%) carried rare, potentially deleterious missense variants in the *LDLR* gene in which pathogenic mutations can cause familial hypercholesterolemia and early-onset cardiovascular disease (*19, 20*). PrimateAI-3D scores of missense variants in *LDLR* were significantly correlated with LDL levels (Spearman ρ = 0.50, P=8×10^−38^) (*16*). Individuals with variants that had scores near 0 had LDL cholesterol levels indistinguishable from non-carriers, whereas those with scores near 1 had elevated LDL cholesterol levels similar to LoF variant carriers (**Fig. 1E**, upper panel). Among individuals who received a clinical diagnosis of dyslipidemia, PrimateAI-3D scores correlated inversely with age of diagnosis (Spearman ρ = −0.35, P = 3×10^−12^). The most deleterious missense variants advanced age of disease onset by ∼15 years, similar to that observed for LoF carriers (**Fig. 1E**, lower panel).

We next examined rare variants in the *PCSK9* gene, a target of cholesterol-lowering medications (*21*). Rare missense variants with high PrimateAI-3D scores in *PCSK9* were correlated with decreased LDL cholesterol levels (Spearman ρ = −0.32, P = 3×10^−13^), and acted in the opposite direction of deleterious *LDLR* variants (**Fig. 1F**, upper panel). LDL cholesterol levels increased with age at a similar rate (0.2 mmol/L per decade of normal aging) regardless of *PCSK9* carrier status, but individuals carrying prioritized rare variants in *PCSK9* had an average of 0.6 mmol/L lower LDL cholesterol levels at any given age (**Fig. 1F**, lower panel). As a consequence, fewer of these carriers had moderate-to-severe hypercholesterolemia (LDL cholesterol > 4.1 mmol/L or 160 mg/dL) or elevated cardiovascular disease risk (*22*), while those that did manifested these symptoms later in life.

Finally, we observed similar relationships between rare deleterious variants in *GCK* and HbA1c, a proxy for blood glucose levels and a diagnostic laboratory marker for type 2 diabetes (pre-diabetes HbA1c > 42 mmol/mol; diabetes HbA1c > 48 mmol/mol) (**Fig. 1G**) (*23*). Analogous to LDL cholesterol, HbA1c levels increased with age, matching the steep rise of diabetes prevalence with age observed in epidemiological studies (*24*). Rare deleterious variants in *GCK* elevated HbA1c levels by an average of 5.1 mmol/mol relative to benign variant carriers and non-carriers, 4.6-fold higher than the average rise in HbA1c levels per decade of normal aging. Correspondingly, this increased the fraction of individuals with diabetes between ages 40-50 from 3.8% to 24.8% (6.6-fold increase) for carriers of rare deleterious variants. In summary, our results across clinically relevant phenotypes such as LDL cholesterol and HbA1c demonstrate the utility of PrimateAI-3D to distinguish pathogenic from benign variants and highlight the capacity of rare high-penetrance variants to accelerate or delay the age of onset of common diseases by decades.

## Rare variant polygenic risk scores identify individuals most at risk for common diseases

Recent exponential human population growth has created an abundance of rare variants via naturally-occurring mutations without providing adequate time for selection to remove those with deleterious consequences (*25, 26*). In the UK Biobank cohort, we observed that each person carries an average of 2.96 rare deleterious missense variants and 0.97 rare LoF variants within one or more of the genes identified from our burden test. Consistent with models of negative selection (*16, 27, 28*), we find that rare variants exerted far greater per allele effects on human phenotypes than common variants across a subset of 893 genes implicated by both rare and common variant studies, with rare deleterious variants having on average an 11.2-fold larger effect than common GWAS variants at the same loci (**Fig. 2A**, **Fig. S4**). Within each allele frequency bin, LoF variants had the highest per allele effects followed by missense variants (PrimateAI-3D > 0.8) and cryptic splice variants (SpliceAI score > 0.2) (*29*). Benign missense (PrimateAI-3D < 0.2) and synonymous variants had nearly null per-allele effects on phenotype, even as singletons. Given the high overall prevalence and strong effect sizes of rare deleterious variants in the predominately healthy UK Biobank cohort, we reasoned that a single polygenic score combining these variants may effectively identify individuals at high risk for complex disease.

**Fig. 2.**
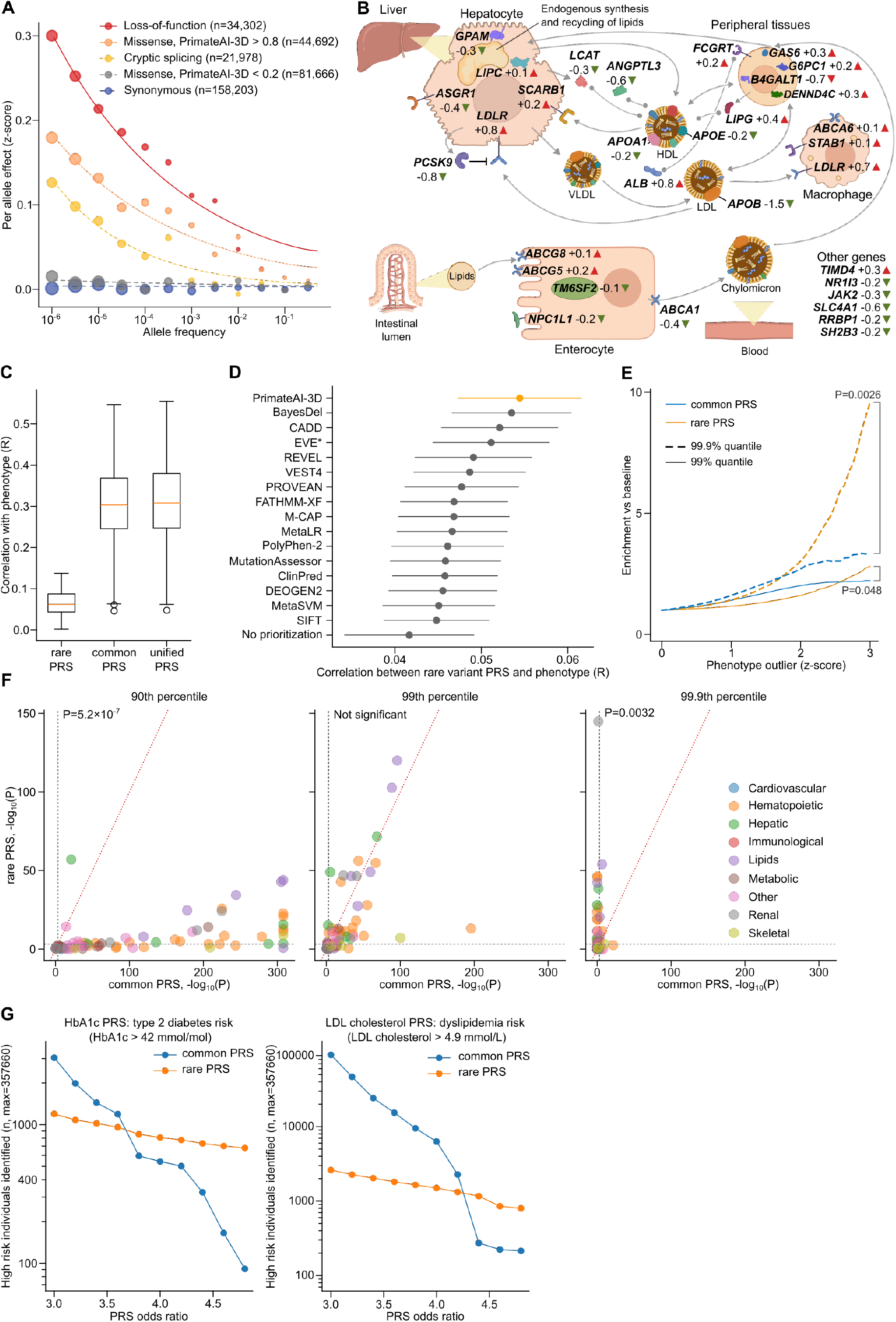
Comparison of polygenic risk scores (PRSs) from common and rare variants. (**A**) Relationship between variant effect size and allele frequency for different pathogenicity classes of variants. Synonymous variants are shown as negative controls. Dot sizes are proportional to the cube root of the number of variants in each group. Regression fits between the allelic effect size and minor allele frequency are shown by curves for each pathogenicity class, calculated using the equation β=*σ*[2*p*(1 – *p*)]^−*a*/2^ where β is the per allele effect, *p* is the minor allele frequency and *σ* and *a* are parameters for selective constraint. (**B**) Illustration of the cholesterol pathway. Genes in the rare variant PRS model are superimposed. For each gene, values indicate effect sizes in standardized units (see Methods), and triangles indicate direction of effect. **(C)** Comparison of the performance of rare variant PRS, common variant PRS, and a unified PRS across 78 phenotypes in the withheld UK Biobank test set. Pearson correlations between PRS predictions and phenotypes are shown. (**D**) Comparison of rare variant PRSs constructed using different pathogenicity classifiers (see Methods). Mean absolute Pearson correlations between PRS and phenotypes are shown. Dots and error bars represent mean ± 95% confidence intervals. **(E)** Enrichment of outlier PRS scores in individuals who are phenotype outliers. Phenotype outlier individuals were defined as exceeding a certain z-score cutoff (x-axis), and the y-axis shows the enrichment of outlier PRS scores in phenotype outlier individuals versus the baseline population, aggregated across 78 phenotypes. **(F)** Comparison of the performance of common variant PRS (x-axis) versus rare variant PRS (y-axis) at identifying individuals at the 90^th^, 99^th^, and 99.9^th^ percentiles (left, middle, and right panels) for 78 quantitative phenotypes. Dashed horizontal and vertical lines show Bonferroni corrected significance thresholds. Lines of equivalence are shown by dashed diagonal red lines. (**G**) Number of individuals at high clinical risk for type 2 diabetes (**left**) and dyslipidemia (**right**), identified by rare and common variant PRSs at varying risk thresholds (x-axis). Rare variant PRSs identified more individuals at higher risk (>3.8 higher odds for type 2 diabetes, and >4.4 higher odds for dyslipidemia) than common variant PRSs.

Existing polygenic risk score (PRS) models of common disease largely omit rare variants due to challenges in interpreting variants of uncertain significance and estimating the magnitude of variant effects (*30*). Here we propose a complementary, rare variant PRS model, based on a weighted sum of rare deleterious variants from multiple phenotype-associated genes, utilizing PrimateAI-3D for variant effect estimation. To construct the model, we first split the UK Biobank cohort into training and testing subsets and then fit a linear model to each phenotype on the rare variants (AF < 0.1%) in associated genes, weighted by PrimateAI-3D predicted effect size (Table S10) (*16*). For comparison, we also constructed common variant (AF >1%) PRS models by performing GWAS on the training dataset and applying the method of clumping and thresholding (Table S11) (*31*).

We illustrate the components of the rare variant PRS model using total cholesterol levels as a representative example and show that it identifies the complex network of genes, cell types, and pathways that underpin lipid metabolism (**Fig. 2B**). Rare deleterious variants in the 31 associated genes that contribute to the rare variant PRS model shifted cholesterol levels by ∼0.38 mmol/L on average, 10-fold the average effect size of the 563 variants in the common variant PRS model (0.040 mmol/L) (Table 1). Out of these 31 genes, 25 were previously known to play central roles in lipid homeostasis (*32*): from absorption of cholesterol via intestinal enterocytes (*ABCG5*) (*33*), to regulation of serum LDL concentrations (*PCSK9*) (*34*), to comprising key components of lipoproteins (*APOB*) (*35*), to lipid scavenging in macrophages (*STAB1*) (*36*). Beyond identifying genes pertinent to cholesterol metabolism, the direction of effect for these rare deleterious variants was consistent with each gene’s known role in the pathway. Notably, many of the genes that produce downregulatory effects on cholesterol levels are therapeutic targets that offer alternatives to statin-based cholesterol reduction for cardiovascular disease, such as *PCSK9* and *NPC1L1* inhibitors (*37, 38*). While the average chance of an individual carrying a rare deleterious variant for any given gene was only 0.4%, when summed across all 31 genes, 1 in 8 individuals carried a rare, high-penetrance variant for cholesterol.

**Table 1:**
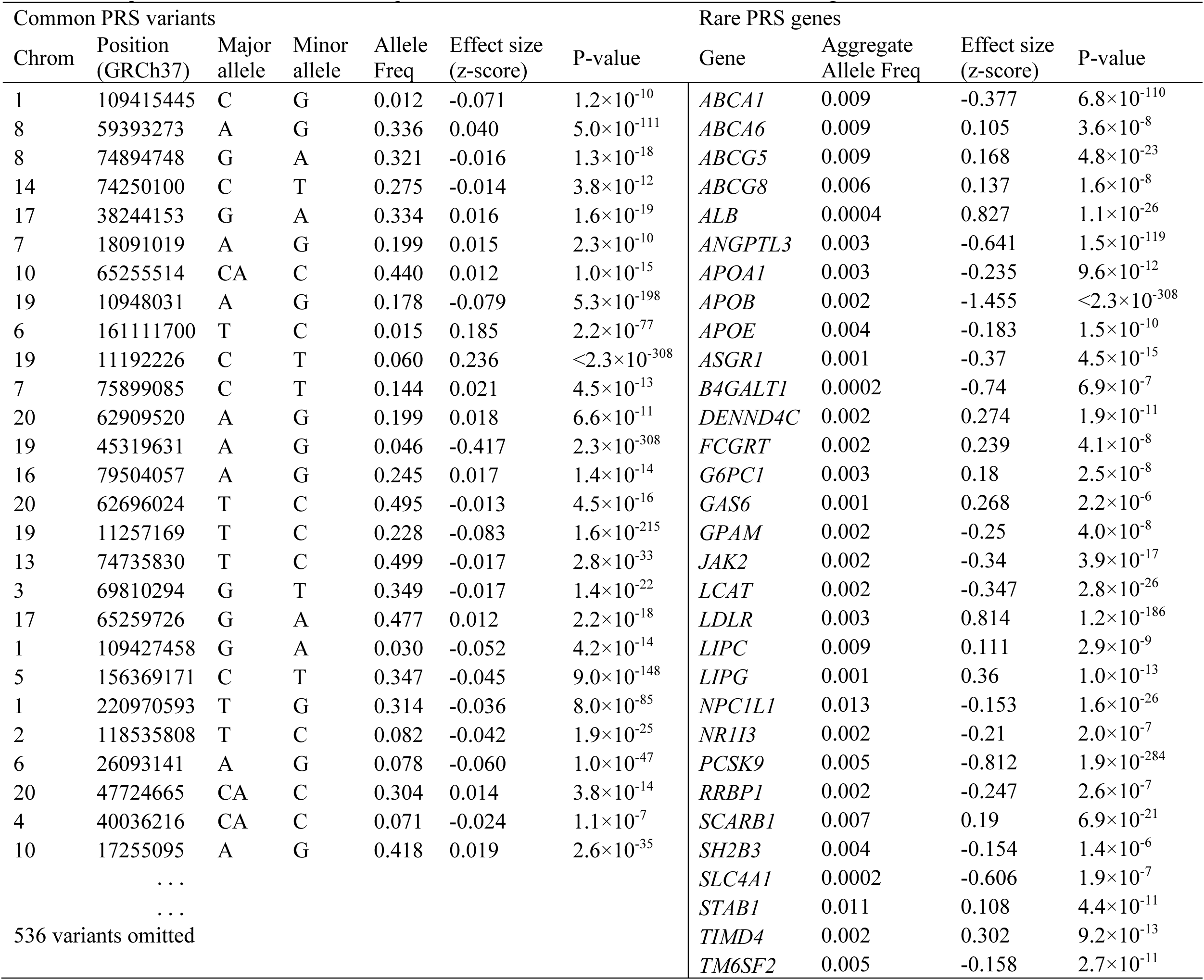
Comparison of effect sizes and frequencies for common PRS variants and rare PRS genes used for normalized cholesterol concentrations. Common PRS variants Rare PRS genes

We sought to evaluate the predictive power of the rare variant PRS and the corresponding common variant PRS, as well as a combination of the two methods, on the 10% of UK Biobank individuals that had been withheld for testing. Across 78 quantitative phenotypes, the unified PRS performed best with an average Pearson correlation of 0.307 (**Fig. 2C**, **Fig. S5**), compared to 0.058 and 0.303 for the rare variant PRS and common variant PRS, respectively. Consistent with the correlations, the average phenotypic variance explained was 10.4%, 0.4%, and 10.1%, respectively. We also evaluated rare variant PRS models constructed using 15 other variant pathogenicity classifiers and observed that PRSs based on PrimateAI-3D outperformed all other methods (**Fig. 2D**), underscoring the importance of accurate pathogenicity prediction to rare variant PRS performance. Overall, these observations are consistent with previous studies that have demonstrated that, in aggregate, rare variants explain less genetic heritability than common variants (*39*).

Although rare variant PRSs underperformed for average phenotype predictions, we reasoned that they may outperform common variant PRSs for identifying individuals at phenotypic extremes, which is more relevant for clinical screening and risk management. Indeed, individuals with an outlier phenotype (z-score ≥3) were 10-fold more likely than the overall population to have a rare variant PRS score in the 0.1^st^ or 99.9^th^ percentile, compared to 3-fold for common variant PRS (P=0.0026, **Fig. 2E**, **Fig. S6**). Across 78 phenotypes, rare variant PRSs significantly outperformed common variant PRSs at identifying individuals with outlier phenotypes at the 99.9% percentile (P=0.0032), had comparable performance at the 99% percentile (difference not significant), and underperformed at the 90% percentile (P=5.2×10^−7^) (**Fig. 2F, Fig. S7)**. Empirically, the prevalence of many complex human diseases is below 1%, including Parkinson’s disease (0.3%) (*40*), multiple sclerosis (0.3%) (*41*), myocardial infarction before age 40 (0.6%) (*42*), and type 1 diabetes (0.2%) (*43*), supporting the relevance of these outlier phenotype thresholds for evaluating clinical risk prediction models.

For two diseases, type 2 diabetes and dyslipidemia, we evaluated the ability of common and rare PRS models to identify individuals exceeding pre-defined diagnostic clinical thresholds (HbA1c > 42 mmol/mol and LDL cholesterol > 4.9 mmol/L, respectively) (**Fig. 2G**). Up until approximately 4-fold increased odds of disease, the common variant PRS identified more at-risk individuals, whereas after this threshold the rare variant PRS overtook the common variant PRS. Because the rare and common variant PRS models use non-overlapping sets of variants, combining them into a unified model enables the identification of significantly more individuals at high disease risk (odds ratio ≥4X) than common variant PRSs alone (type 2 diabetes, 1912 vs 542, P=1.4×10^−178^, dyslipidemia 7858 vs 6306, P=1.2×10^−39^). Taken together, these findings suggest that incorporating rare variants into PRSs can outperform common variant PRSs for identifying outlier individuals (*30, 44*) who are most likely to require treatment or to suffer severe, early-onset manifestations of disease and for whom preventative screening would be most impactful (*45, 46*). Moreover, the ability to point to a single penetrant variant as the primary cause of the phenotype may increase the potential clinical actionability of rare deleterious variants with respect to prognosis, management, and therapeutic interventions (*47*).

## Portability of rare variant polygenic risk scores, and validation in an independent, multi-ancestry cohort

Common variant PRS models derived from European populations have poor portability in non-European populations, which may contribute to future health disparities once adopted into clinical practice (*4*). Even when applied to populations with similar ancestry, common variant PRSs have decreased performance owing to differences between the cohorts used for training and testing (*48, 49*). We thus set out to evaluate the robustness of our rare variant PRSs across independent cohorts and ancestries. We first applied 16 rare variant PRS models, which had been trained on UK Biobank European-ancestry individuals, to predict quantitative phenotypes in 20,708 European individuals from the Massachusetts General Brigham Biobank (MGB, Table S12) (*50*). Across 16 phenotypes, the average predictive performance of the rare variant PRS model was similar in the two cohorts (Pearson R = 0.53), with a median phenotype correlation of 0.078 between the rare variant PRS and the UK Biobank withheld test cohort, compared to 0.084 for the MGB cohort (**Fig. 3A**). Notably, the rare variant PRS models achieve approximately equal performance in the two cohorts despite 43% of the rare deleterious variants in the MGB cohort never appearing in the UK Biobank cohort that was used for model training. Thus, unlike common variant PRSs, rare variant PRSs appear largely portable across cohorts with similar ancestry.

**Fig. 3.**
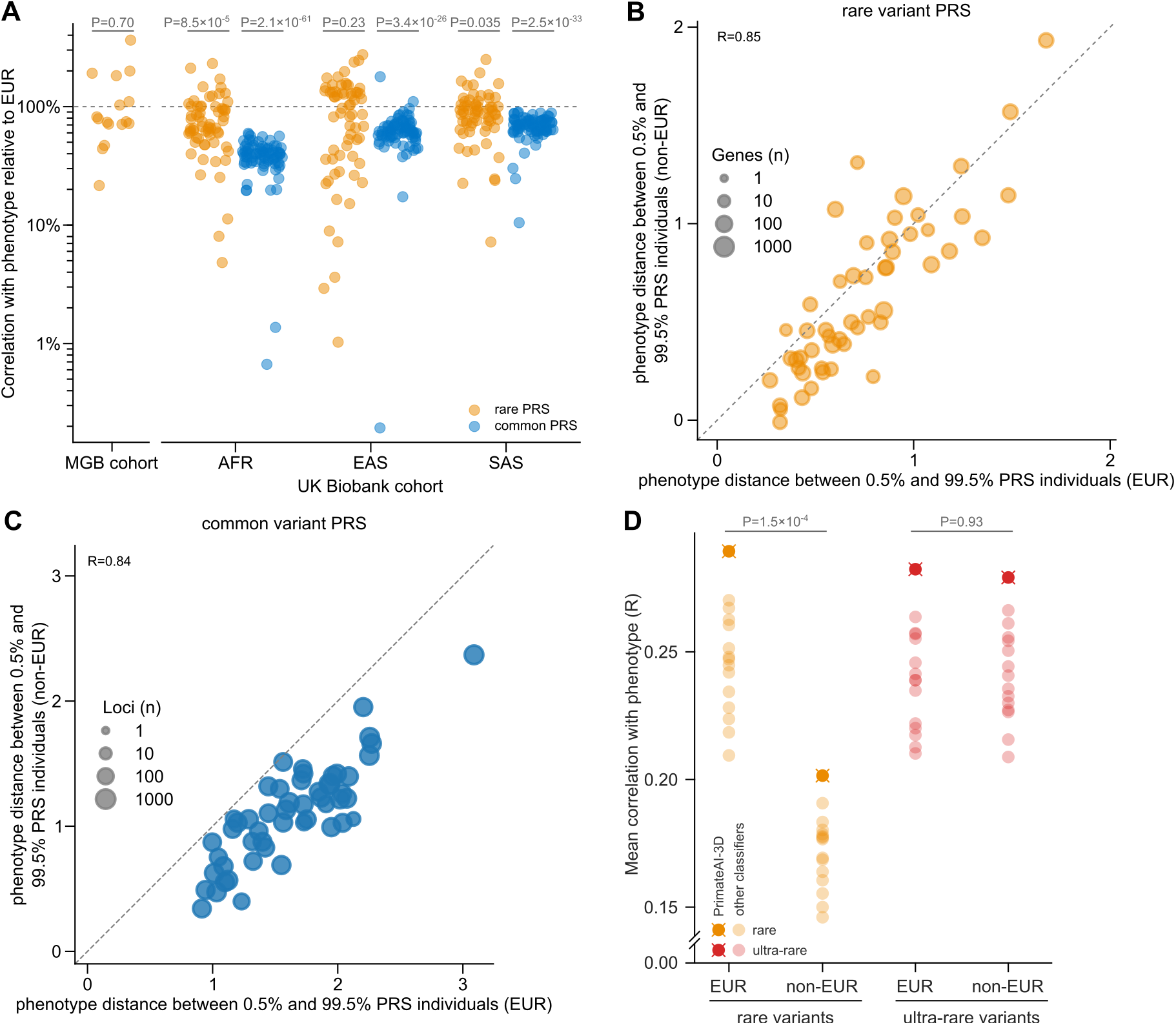
Validation of rare variant PRS performance in diverse human populations. (**A**) Performance of rare and common variant PRSs derived from UK Biobank Europeans (EUR), measured in the MGB cohort (left) and in UK Biobank non-Europeans (non-EUR) stratified by ancestry (right, AFR: African, EAS: East Asian, SAS: South Asian). Performance is shown relative to held out European individuals in the UK Biobank. P-values indicate whether the difference in performance versus held out Europeans is significant. (**B**) Mean phenotype distance between UK Biobank EUR (x-axis) and UK Biobank non-EUR (y-axis) individuals is shown for 52 matching traits. The phenotypic distance is calculated by comparing individuals with low (<0.5%) and high (>99.5%) rare variant PRS percentiles. The Pearson correlation is reported. A line of equivalence is shown by the gray diagonal dashed line. (**C**) Same as (B) but showing the results for common variant PRSs. (**D**) Performance of PrimateAI-3D variant effect predictions stratified by ancestry and allele frequency for 49 gene-phenotype pairs. Correlation of predicted variant effects with observed phenotypes is shown on the y-axis. Rare variants have AF < 0.1% in each population. Ultra-rare variants are absent from TOPMed, and non-EUR ultra-rare variants are singletons (AC=1) whereas EUR ultra-rare variants have allele frequencies less than or equal to those of the non-EUR singletons. P-values are displayed for comparisons across ancestries using PrimateAI-3D. The performance of other variant classifiers is also shown for context.

We next evaluated the performance of our rare and common variant PRS models, which had been trained only on individuals of European ancestry, in individuals of non-European ancestry from the UK Biobank and MGB. As a control, we ensured that the number of variants used per person in the rare variant PRS was closely matched for different ancestries by applying ancestry-specific allele frequency filters (AF < 0.1%) (**Fig. S8**) and verified that the resulting PRS distributions were similar across ancestries (**Fig. S9**). Consistent with previous reports, the median common variant PRS correlation with phenotype was 84% lower in individuals with African ancestry (P=2.1×10^−61^), 62% lower in individuals with East Asian ancestry (P=3.4×10^−26^), and 51% lower in individuals with South Asian ancestry (P=2.5×10^−33^) relative to the correlation in individuals with European ancestry (**Fig. 3A**). In contrast, the rare variant PRS correlation was substantially more portable with smaller reductions in median correlation of 54%, 14%, and 23%, respectively. To assess the portability of the rare variant PRS on a more clinically relevant task, we selected individuals with PRS scores at the upper and lower ends of the phenotype distribution (top or bottom 0.5%) and observed that the average phenotype differences between the two groups were similar for Europeans and non-Europeans in both the UK Biobank withheld test cohort (Pearson R = 0.85, **Fig. 3B**) and the MGB cohort (Pearson R = 0.88, **Fig. S10**). Overall, rare variant PRS models trained in Europeans performed better when tested in non-Europeans than Europeans for 14 out of 52 phenotypes, compared to the common variant PRS models which performed worse when tested in non-Europeans for all 52 phenotypes (**Fig. 3C**).

While rare variant PRSs appear to generalize better across ancestries than common variant PRSs, their average performance still decreases in non-European populations. However, this appears to be distinct from the portability issues experienced by the common variant PRS, where causal variant identification remains difficult due to linkage disequilibrium. We hypothesized that the current European bias is due primarily to more accurate allele frequency estimates within the more numerous European individuals in the cohort and in current population databases, resulting in the inadvertent inclusion of common non-European variants into the rare variant PRS that dilute its performance. To test this hypothesis, we restricted our evaluation to ultra-rare variants (seen only once in the UK Biobank and absent from the TOPMed allele frequency database) to minimize common variant leakage. We found that PrimateAI-3D variant effect size predictions were equally accurate in European and non-European ultra-rare variants (difference not significant, **Fig. 3D**) but were significantly less accurate for non-European variants at the default allele frequency threshold of 0.1% (P=1.5×10^−4^ using PrimateAI-3D). As further indication that these issues are independent of variant effect prediction, we show that rare variant PRSs derived using only LoF variants (without PrimateAI-3D) displayed similarly decreased performance in non-European individuals (**Fig. S11A**) and that the European bias could be reduced by using L1 regularization to limit overfitting (**Fig. S11B**). Similar challenges have been reported for rare genetic disease diagnosis in non-European populations (*51, 52*), where inaccurate allele frequency estimates make it difficult to preclude ancestry-specific common variants as potential causes of disease. Therefore, as population allele frequency panels become more accurate and globally inclusive, we expect that the portability of rare variant PRSs will continue to improve.

## The convergence of common and rare variant genes forecasts future improvements in rare variant PRSs

Looking forward, we explored how much the performance of rare variant PRS approaches is expected to improve as exome sample sizes increase, focusing first on our ability to identify additional exome-wide significant genes (FDR < 5%). We performed association tests in down-sampled subsets of the UK Biobank cohort and observed that the number of significant associations increased linearly with sample size for both rare variant burden tests (FDR < 5%) and common variant GWAS loci (P < 5×10^−8^) (**Fig. 4A, Fig. S12**). On average, PrimateAI-3D enabled discovery of the same number of exome-wide significant genes using 1.8-fold smaller cohort sizes compared to when missense prioritization was not applied. Consistent with the improved detection of phenotype-associated genes, we observed a linear increase in the number of variants carried by each individual that could be included in the rare variant PRS model (**Fig. 4B**). At the full cohort size, we found that 97% of individuals carried a rare penetrant variant in one or more of the associated genes for the 90 clinical and quantitative phenotypes in the study (**Fig. S13**). Although effect sizes were lower in newly identified genes (**Fig. 4C**), rare variant PRS performance improved steadily, with each doubling of discovery cohort size corresponding to an 88% improvement in variance explained (**Fig. 4D, Fig. S14**).

**Fig. 4.**
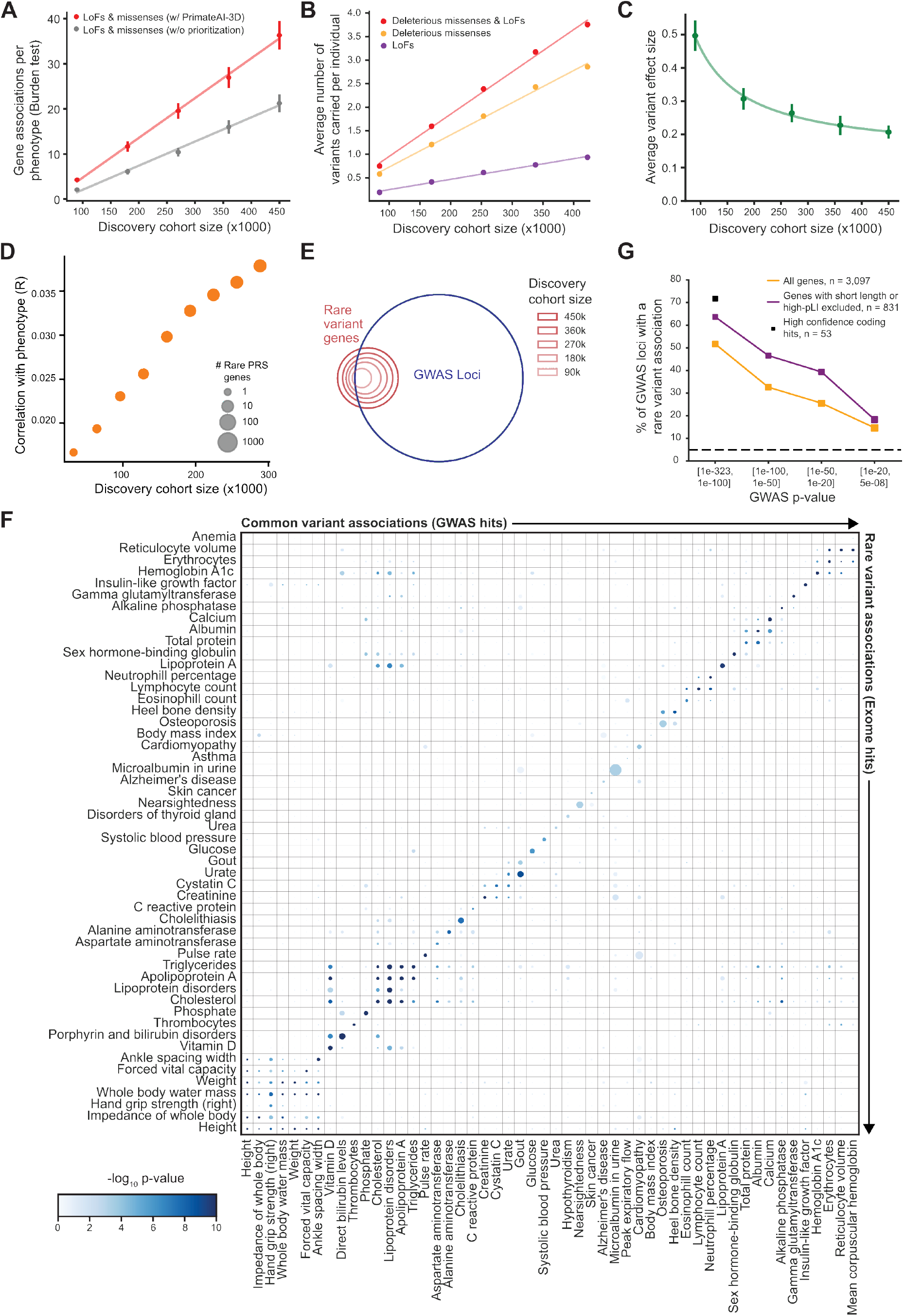
Forecasting the growth of rare variant associations with increasing cohort size. (**A**) Number of significant (FDR < 0.05) genes identified per phenotype with rare variant burden tests as a function of the discovery cohort size in thousands of individuals. Missense prioritization with PrimateAI-3D substantially increased the number of genes detected at all cohort sizes. Dots and bars represent mean ± standard error. (**B**) Number of rare deleterious variants identified per individual as a function of the discovery cohort size. (**C**) Average per variant absolute effect size for newly associated genes (FDR < 0.05) at each discovery cohort size. The fit from the regression *y* = *a/x* + *b* is shown. Dots and error bars represent mean ± standard error. **(D)** Rare variant PRS performance increases with increasing discovery cohort size. Median correlation between the PRSs and the phenotype is shown on the y-axis. Number of genes included in the PRS is represented by the size of each point. **(E)** Venn diagram showing the overlap of rare variant genes with common variant GWAS loci as a function of discovery cohort size. **(F)** A non-symmetrical heatmap showing the phenotype-specific overlap of common and rare variant associations. Each point shows the statistical significance of the overlap between common variant GWAS genes associated with the x-axis phenotype and rare variant genes associated with the y-axis phenotype. The size of the points represents the magnitude of the enrichment while the color represents the P-value. **(G)** Percentage of unambiguously mapped GWAS genes with rare variant associations (nominal P-value ≤ 0.05) stratified by GWAS significance thresholds. Results are shown for all genes (orange) and after excluding genes that are less likely to show rare variant signal (purple) due to short length (<2 kb coding sequence) or strong selective constraint (pLI > 0.99, probability of being loss-of-function intolerant). High-confidence coding hits are defined as having a lead variant with GWAS P < 10^−100^ with strong linkage disequilibrium (r^2^ >= 0.9) to a coding variant in the associated gene. The dashed line shows the background false discovery rate.

Our forecasting analyses suggest that rare variant PRSs will continue to meaningfully improve as cohort sizes increase, with newly discovered genes preferentially enriched at GWAS loci (**Fig. 4E**), consistent with recent work showing convergent biological pathways behind both rare and common variant heritability (*39*). The observed overlap of common variant GWAS hits and rare variant burden test genes was highly phenotype-specific (**Fig. 4F, Fig. S15, Fig. S16**), and was not explained by linkage disequilibrium, as we regressed out the effects of significant GWAS variants and population structure before applying the rare variant burden tests. Focusing on a subset of well-powered GWAS loci that could be unambiguously mapped to a single protein-coding gene (*16*), we found that 64% of common variant GWAS genes showed significant association in the rare variant burden test (P < 0.05, **Fig. 4G**). The fraction of genes with rare variant signal declined for weaker GWAS hits (P = 3×10^−37^), as well as for genes under strong evolutionary selection (P = 5×10^−4^) (*53*), reflecting reduced statistical power to detect enrichments in genes that either have weak phenotypic effects, or that have been depleted of deleterious variants by selective constraint. Similarly, we observed that shorter genes, with consequently fewer variants, were also less likely to be significant in the rare variant burden test (P = 7×10^−6^). Although we found that only 186 (6%) out of 3,097 unambiguously GWAS-implicated genes reached the stringent exome-wide significance threshold for inclusion in the rare variant PRS (FDR < 5%), 625 (20%) were nominally significant on the burden test at a P-value threshold < 0.05, indicating that rare variant associations are likely to be discovered at these genes with larger cohort sizes. In summary, our empirical studies of the convergence of common and rare variant associations suggest that allelic series underlie most of the genes implicated in human pathophysiology and can be leveraged in ever growing sequencing cohorts to improve rare variant PRS performance.

## Discussion

Understanding the role of rare penetrant variants in common diseases is of prime interest to both precision medicine (*5–7*) and targeted drug development (*21, 54, 55*). In this study, we leverage PrimateAI-3D’s state-of-the-art predictions to model the quantitative effects of each variant on multiple phenotypes, uncovering the role played by rare penetrant variants in common human diseases and complex traits. We demonstrate the complementary utility of common and rare variants for predicting the risk of human diseases, observing that common variants explain a higher proportion of total population variance, whereas rare variants more readily identify outlier individuals at the greatest risk for severe, early-onset disease (*45, 46*). Our results establish that the personal genome of an otherwise healthy individual is not quiescent with limited actionable potential (*56*) but instead carries a substantial burden of rare consequential variants, the clinical utility of which will be more fully realized as variant interpretation improves and discovery cohort sizes increase.

At present, the two greatest barriers to the clinical adoption of common variant PRS models for use in precision medicine are their limited generalizability between populations with different ancestries and their weak discriminatory capability to identify individuals at high risk for disease (*57*). Specifically, the inclusion of predominately non-coding variants with small effects that are non-causal, but disease-associated due to linkage disequilibrium, substantially impairs common variant PRS performance (*58, 59*). In comparison, our rare variant PRS models are anchored on PrimateAI-3D’s predictions of missense variant effect size and are largely uninfluenced by the effects of ancestry, since the PrimateAI-3D model was derived from common variants in 236 species of non-human primates. This gives rare variant PRS models an advantage over common variant PRS models at generalizing to cohorts and human populations that were not seen during training, providing more globally equitable health outcomes than current genetic studies that are predominantly European. Ultimately, rare variant PRSs can be combined with common variant PRSs into a unified risk model to significantly improve the identification of individuals from the general population who are at increased risk for common diseases.

Although the rare variant PRS models presented in this work show promise for accurate identification of high-risk individuals across diverse human populations, our study has several limitations. At present, rare variant PRS models have limited power; we are only capable of robustly estimating variant effects for well-powered genes, finding 217 GWAS loci but only 34 rare variant genes on average per trait. We empirically forecast that the exact causal genes underlying most of these GWAS loci will be uncovered by rare variant studies with larger cohort sizes and advances in variant interpretation algorithms (*60*). Second, while interpretation of variants of uncertain significance (VUS) remains a challenge, recent advances applying deep learning (*12, 61*), high-throughput experimental assays (*62*), and variant information from closely related primate species (*63*) have each demonstrated promise towards solving variant interpretation on a genome-wide scale. Third, although we observed improved portability across ancestries for rare variant polygenic prediction, more accurate allele frequency resources for global populations will further shrink the discrepancies in performance across populations. Indeed, systematic efforts to catalog rare variation in non-European populations are ongoing (*64, 65*) and will likely precede well-powered common variant GWAS studies in diverse global populations (*66*). Finally, although we only evaluate rare coding or splice altering variants, improved non-coding variant prediction coupled with larger sample sizes would likely reveal the pervasive phenotypic impacts of rare penetrant variants in each person, with transformative implications for the utility of clinical whole-genome sequencing in the general population.

## Methods summary

### Datasets

We analyzed data from unrelated individuals in the UK Biobank, all of whom had genotypes obtained from microarrays and 454,712 of whom had genotypes available from exome-sequencing. The work described in this manuscript was approved by the UK Biobank under application no. 33751. In addition, we performed validation experiments with 20,708 individuals from the MGB Biobank.

### Phenotype processing

Quantitative traits were standardized by inverse rank normal-transformation and adjusted for medication usage and further covariates including age, sex, ancestry, diet and others. Binary traits were adjusted for age, age^2^, sex, age × sex, age^2^ × sex and ancestry.

### Common variant associations

GWAS were performed with common variants (AF > 1%) in individuals of European ancestry in the UK Biobank and causal gene sets were derived by linkage disequilibrium between independent GWAS significant variants (P < 5×10^−8^) and coding variants, splicing variants or eQTLs in nearby genes or by proximity with local transcription start sites.

### Rare variant associations

Burden tests were performed with rare variants (AF < 0.1%) on individuals from all ethnicities by searching for combinations of allele frequencies and missense pathogenicity scores per gene and further calibrating via permutations to maximize significance prior to FDR correction. Significant gene-phenotype pairs were reported at 5% FDR after correction for multiple hypothesis testing across all autosomal protein coding genes in the human genome and across all tested traits. Rare variant results generated in this study were compared to results from a recent well-powered rare variant analysis in the UK Biobank (*17*) by examining the overlap of significant genes, along with the enrichment of GWAS and clinically relevant genes. Multiple missense classifiers were considered for pathogenicity prediction in the burden tests, including BayesDel (*67*), CADD (*68*), ClinPred (*69*), DEOGEN2, EVE* (*61*), FATHMM-XF (*70*), M-CAP (*71*), MetaLR (*72*), MetaSVM (*72*), MutationAssessor (*73*), Polyphen-2 (*74*), PrimateAI-3D (*12*), PROVEAN (*75*), REVEL (*76*), SIFT (*77*), and VEST4 (*78*). Scores for the EVE-style variational autoencoder (EVE*) were generated by reimplementing the method. The different classifiers were compared via Spearman correlation with the average phenotype values of the carriers of each qualifying missense variant in high-confidence associated gene-phenotype pairs.

### Polygenic risk scores

PRS models were constructed from GWAS and burden test results from training datasets. Common variant (AF >1%) PRS models were constructed by applying the method of clumping and thresholding (*31*). In contrast, rare variant PRS models were constructed by fitting linear models to each phenotype on the rare variants (AF < 0.1%) in significantly associated genes, weighted by predicted missense pathogenicity. A unified PRS model was also constructed, which summed the rare and common variant PRS models per individual. As with the burden test results, rare variant PRS performance was evaluated using PrimateAI-3D and other classifiers across 78 traits. The overlap of individuals at phenotypic and PRS extremes was examined to further elucidate PRS performance. For two traits, HbA1c and LDL cholesterol, clinical risk prediction was assessed, since clinically diagnostic thresholds could distinguish cases from controls. PRS portability was assessed in two ways - firstly between cohorts, by applying models constructed in the UK Biobank to the MGB Biobank, and secondly between ancestries, by comparing the performance between different ancestry groups in the UK Biobank.

### Forecasting analysis

Growth projections of rare and common variant associations, PRS performance, and overlap of significantly associated genes from rare and common variants were made from randomly down-sampled data ranging from 20% to 100% of the whole UK Biobank exome cohort with 20% increments.

Full materials and methods are available in the supplementary materials (*16*).

## Data Availability

PrimateAI-3D prediction scores are available with a noncommercial license upon request and are displayed at https://primad.basespace.illumina.com. 
Source code is available open source under an academic license upon request

https://github.com/Illumina/PrimateAI-3D

## Acknowledgments

We would like to thank Daniel MacArthur, Jonathan Pritchard, Manuel Rivas, Nicole Ersaro, and Ileena Mitra for helpful discussions, and the participants and investigators in the UK Biobank (Resource Application Number 33751) and MGB studies (protocol 2018P001236) who made this work possible.

## Author contributions

PF, JM, JU, JD, TH, YY, PW, ZN, JS, HG, AM, DC, FA, and KF performed the analysis and wrote the manuscript. JR, TMB, MR, HR, AOL, AK, and KF supervised the work.

## Data and materials availability

PrimateAI-3D prediction scores are available with a non-commercial license upon request and are displayed at https://primad.basespace.illumina.com. Source code is available open source under an academic license upon request.

## Supplementary Materials

Materials and Methods

Figs. S1 to S16

Supplemental Tables S1 to S12

References (79–*88*)

