## Supplemental Methods and Figures for "Rare penetrant mutations confer severe risk of common diseases"

**This file includes:**

Materials and Methods

Figs. S1 to S16

**Other Supplementary Materials for this manuscript include the following:**

Supplemental Tables S1 to S12

### Materials and Methods

#### Cohorts

##### GWAS cohort

We used a total of 357,661 unrelated individuals of self-declared white British ancestry together with a total of 13,791,468 genotype calls for common variants, which were either directly measured or imputed. In brief, individuals were restricted to white British genetic ancestry with exclusion of closely related individuals (or at least one of a related pair of individuals), individuals with sex chromosome aneuploidies, and individuals who had withdrawn consent from the UK Biobank study. The full quality control procedure for individuals and genotypes is described here (*79*).

##### Exome cohort

For our exome sequencing cohorts, we used data from whole exome sequencing of 454,712 individuals released by the UK Biobank in October 2021. To reduce confounding by cryptic relatedness, we excluded individuals who have more than second degree relatedness based on their kinship coefficients as provided by the UK Biobank. For each pair of individuals with a kinship coefficient greater or equal to 0.0884, we kept one of the individuals at random and excluded the other one. To maximize power in our burden tests, we kept individuals from all ethnicities. After quality control, our exome cohort consisted of 423,614 total individuals. However, cohorts for different phenotypes could be smaller, because not every phenotype was available for every individual with exome sequencing data (Table S1). We also conducted validation experiments with the Mass General Brigham Biobank (MGB Biobank), containing 20,708 individuals with exome-based genotypes. The Massachusetts General Hospital IRB approved the present secondary data analysis under protocol 2018P001236.

#### Phenotypes

##### Quantitative phenotypes

We used quantitative anthropometric (e.g. height, weight, body fat percentage etc) phenotypes, blood biomarkers (e.g. glucose and LDL concentrations), urine biomarkers (e.g. Creatinine) and Spherical equivalent (which is a quantitative measure for nearsightedness). To be sufficiently powered, we required that each quantitative phenotype is measured in at least 100,000 individuals out of the whole UK Biobank cohort. We further excluded Testosterone, which is a highly sex specific phenotype. For systolic and diastolic blood pressure, we took the average of the two consecutive measurements at each visit. For Spherical Equivalent, we first computed the values for the left and the right eyes as “Spherical power” + 0.5 × “Cylindrical power” and then took the average between the two eyes as the phenotype value for each person.

Each phenotype was corrected for drug usage (e.g. statins, blood pressure medications, see next section and Table S2), then inverse-rank normal transformed and further corrected for a number of other covariates (e.g. age, sex, genetic principal components, diet, smoking status, see Table S3). Quantitative phenotypes were further pruned to a non-redundant set so that every pair of phenotypes had an absolute pairwise Pearson correlation of at most 0.95. Among each group of redundant phenotypes, the phenotype with the most individuals was selected.

##### Correction for drug use

Each quantitative phenotype was corrected for drug use by applying the following procedure. First, drugs were grouped together in 34 categories by using the grouping in (*80*). Effects of different drug classes on each quantitative phenotype were estimated from a subset of the participants (approximately 20,000 individuals), who came for a repeated visit (t2) approximately 5 years after the initial visit (t1) and had their quantitative phenotypes and corresponding covariates recorded again, including current drug use. For each quantitative phenotype $Y$, covariate-corrected values at time point t1 and t2 ($\tilde{Y}_{t1}$ and $\tilde{Y}_{t2}$) were computed by regressing out all covariates (Table S3) except drug use and the difference between the two time points was computed as:

$$\delta= \tilde{Y}_{t2}- \tilde{Y}_{t1}$$

Then, for each drug category X, all UK Biobank individuals were partitioned into four groups according to their drug use and a binary indicator variable was introduced for each group that encodes whether an individual belongs to that group:

1) Individuals who were not taking drug X at both time points, t1 and t2 (indicator variable: $X_{00})$
2) Individuals who had started taking drug X between time point t1 and t2 (indicator variable: $X_{01})$
3) Individuals who had stopped taking drug X between time point t1 and t2 (indicator variable: $X_{10})$
4) Individuals who were taking drug X at both time points, t1 and t2 (indicator variable: $X_{11})$

To determine which drugs have a significant effect on the phenotype, a forward selection stepwise regression of the form below was fitted iterating over all drug categories:

$$\delta= \beta_{00}X_{00}+ \beta_{01}X_{01}+ \beta_{10}X_{10}+ \beta_{11}X_{11}+ \beta_{t}t+ \beta_{m}\left( \tilde{Y}_{t1}- mean\left( \tilde{Y}_{t1} \right) \right)+\beta_{0}$$

In the above equation, the term $\beta_{t}t$ models the effect of the time passed between t1 and t2 (t = t2 – t1 for each individual), the term $\beta_{m}\left( \tilde{Y}_{t1}- mean\left( \tilde{Y}_{t1} \right) \right)$ accounts for effects due to regression to the mean between the two time points, and $\beta_{0}$ is the intercept (*81*). At each step, the most significant drug was selected based on the P-values of the $\beta_{01}$ coefficients and was included in the current set of significantly associated drugs, $D'$, which in turn was regressed out in the subsequent steps with respect to all drug use patterns, $P=\{00, 01, 10, 11\}$, in the following regression:

$$\delta= \beta_{00}X_{00}+\beta_{01}X_{01}+\beta_{10}X_{10}+\beta_{11}X_{11}+\beta_{t}t+\sum_{d\in D'} \sum_{p\in P} \beta_{p}^{(d)}X_{p}^{(d)}+\beta_{m}\left( \tilde{Y}_{t1}- mean\left( \tilde{Y}_{t1} \right) \right)+\beta_{0}$$

The stepwise procedure was stopped when the P-value of the coefficient $\beta_{01}$, which models the effect of starting drug X, had increased above 10^-3^.

After the set of relevant drugs ($D)$ was determined, their individual effects were estimated jointly by fitting the following regression:

$$\delta= \sum_{d\in D} (\beta_{00}^{\left( d \right)}X_{00}^{\left( d \right)}+ \beta_{01}^{\left( d \right)}X_{01}^{\left( d \right)}+\beta_{10}^{(d)}X_{10}^{(d)}+ \beta_{11}^{(d)}X_{11}^{(d)})+ \beta_{t}t+ \beta_{m}\left( \tilde{Y}_{t1}- mean\left( \tilde{Y}_{t1} \right) \right)+\beta_{0}$$

The raw values for phenotype Y at time point t1 across all individual in the UK Biobank including those who were not present at the repeated visit were then corrected as:

$$\hat{Y}_{t1}=Y_{t1}-\sum_{d\in D} \left( \beta_{01}^{\left( d \right)}- \beta_{00}^{\left( d \right)} \right)X^{\left( d \right)}$$

Where $X^{(d)}$ is a binary indicator variable that encodes whether an individual was taking drug *X* at the initial visit, t1. After the correction for drug use, the values $\hat{Y}_{t1}$ were inverse-rank normal transformed and all other covariates (Table S3) were regressed out. The values obtained in this way were used as input for all further analysis in the paper unless in Figs. 1F and 1G.

To show the relationship between age and phenotype for carriers of pathogenic and benign missense variants in Figs. 1F and 1G, we applied the above steps for covariate and drug use correction except that we did not apply inverse rank normal transformation before regressing out all other covariates. Subsequently, we added back the effects of “age”, “age^2^”, “age × sex” and “age^2^ × sex”, then regressed out “sex” and, finally, added back the mean of the original phenotype values. In this way, we retained the original scale, which is meaningful for clinical interpretation.

For a small number of quantitative phenotypes, the phenotype was not measured at the second visit: Ankle spacing width, Heel bone mineral density BMD, Heel bone mineral density BMD T score automated, Heel Broadband ultrasound attenuation direct entry, Heel quantitative ultrasound index QUI direct entry and Speed of sound through heel. To correct these phenotypes for drug use, drug use at the initial visit was included as a binary indicator variable together with the rest of the covariates.

##### Clinical phenotypes

Clinical phenotypes were derived as binary variables from the category of “First occurrences – Health-related outcomes” (Category 1712) and were set to 1 if a value was present in the corresponding “Date XX first reported” (where XX stands for the ICD10 code for the disease), and 0 otherwise. Age at diagnosis was derived for each clinical phenotype as the difference between the values in the corresponding “Date XX first reported” and the birth date of each individual.

#### Rare variant analyses

##### Rare variant principal components

To compute rare-variants principal components (PC) we first identified 610,063 variants with 0.00005 < AF < 0.01 in 454,712 individuals with whole-exome sequence data available using plink 1.9 (*82*). We then selected 420,238 independent variants (window size of 2000 variants and r^2^ threshold < 0.01) across all autosomes. We finally used a fast-principal component (*83*) as implemented in plink 2.0 (*82*) to calculate 25 PCs, of which we selected the first 20 eigenvectors.

##### PrimateAI-3D scores for missense variants

Raw PrimateAI-3D scores were converted to percentiles within each gene. The PrimateAI-3D percentiles are referred to as “PrimateAI-3D scores” in our study and were used in all downstream analyses as a measure of pathogenicity for each missense variant in the UK Biobank 450k exome cohort. Downstream analyses were performed with rank-based statistics (Spearman correlation, etc) so that results are independent of such monotonic transformations of the scores.

##### Rare variant burden tests

We tested for association between traits and presence of rare variants per gene. Variants from the UK Biobank 450k exome release were annotated with VEP v105.0 to obtain predicted variant consequences, along with HGNC symbols from GENCODE v39 and gnomAD allele frequencies. We excluded variants with allele frequency greater than 0.001 in either the UK Biobank exome cohort or the population with highest allele frequency in gnomAD (popmax AF), or variants for which more than 5% of individuals had missing genotype calls.

Our tests included loss-of-function variants (LoF: stop_gained, stop_lost, frameshift_variant, splice_donor_variant, splice_acceptor_variant, start_lost, transcript_ablation, transcript_truncation, exon_loss_variant, gene_fusion, bidirectional_gene_fusion, or SpliceAI > 0.2), missense variants and synonymous variants (as negative controls). For each gene, we partitioned individuals in the exome cohort into carriers and non-carriers of rare variants across multiple consequence categories (LoF, missense-only, deleterious (LoF or missense with high predicted pathogenicity), all LoFs and missenses without prioritization and synonymous) and tested if the phenotype values between carriers and non-carriers were significantly different. For quantitative phenotypes, we first regressed out independent genome-wide significant common GWAS variants (P < 5×10^-8^) from the phenotype (after adjustments for drug usage and covariates described earlier), along with 20 rare variant principal components and applied a two-tailed t-test. For binary phenotypes, we fitted a logistic regression with carrier status as a binary indicator variable and age, sex, all 40 common variant genetic principal components and 20 rare variant genetic principal components, genome-wide significant common GWAS variants and a bias term as covariates.

To maximize power, for each gene we performed a grid search over the allele counts and predicted pathogenicity scores of the observed variants to find the cutoffs for maximum allele count and minimum predicted pathogenicity score that maximizes the significance of the burden test. The grid search across predicted pathogenicity scores limited to five values for PrimateAI-3D scores, corresponding to the five quantiles of the observed scores for the gene. Thus, for each gene the number of tests performed was N_AC_ × 5, where N_AC_ was the number of unique allele counts across variants observed in the gene for the consequence category in the exome cohort.

To account for the multiple testing burden due to these grid searches for each gene, we performed two types of multiple testing correction: Benjamini-Hochberg False Discovery Rate (BH FDR) correction and permutation tests. BH FDR was more computationally efficient but overcorrected for multiple testing within the same gene. For this reason, we used BH FDR correction only for genes where the BH FDR corrected P-value was less than 10^-5^. In genes with BH FDR-corrected P > 10^-5^, which was most gene-phenotype tests, we used adaptive permutation tests to estimate the true P-value, using up to 100,000 randomizations of the phenotype values per gene. To reduce the computational demand, the number of permutations for each gene was scaled so that genes with less significant uncorrected P-values received fewer permutations. Specifically, the number of permutations for each gene was determined as the lesser of 100/P-value and 100,000.

For binary traits, fitting logistic regressions on the permuted data was not computationally feasible. Instead, we used chi-squared tests without covariate adjustment as a proxy to estimate the P-value inflation caused by the grid search. We obtained a P-value ($P_{raw, \chi2}$) using a chi-squared test with the thresholds for allele counts and PrimateAI-3D determined from the optimal fitted logistic model ($P_{raw, Logit}$), and corrected the chi-squared P-value by chi-squared permutation tests ($P_{adj, \chi2}$). We then adjusted the P-value of the fitted logistic model as:

$$\gamma= \frac{Log(1-P_{adj, \chi2})}{Log(1-P_{raw, \chi2})}$$

$$P_{adj, Logit}=1-{(1-P_{raw, Logit})}^{\gamma}$$

In the above equations, $\gamma$ denotes the inflation bias determined by the permutation tests using chi-squared tests. $P_{adj, Logit}$ denotes the corrected P-value from the logistic model after accounting for the inflation bias, $\gamma$. Finally, to correct for potential global deviations from the null distribution of the adjusted P-values from the burden test for binary phenotypes, $P_{adj, Logit}$, we applied genomic correction on those P-values by computing:

$$\lambda=-\frac{{Log}_{10}(2)}{{Log}_{10}(1-P_{raw, Logit, m})}$$

$$\hat{P}=1-{(1-P_{raw, Logit})}^{\gamma*\frac{\lambda}{\gamma_{m}}}$$

where $P_{raw, Logit, m}$ and $\gamma_{m}$ denote the median $P_{adj, Logit}$ value and the inflation bias of the gene with the median $P_{adj, Logit}$ value, respectively, across all tested genes in the human genome. We used $\hat{P}$ as the final P-value for each gene from the burden test for binary phenotypes.

For quantitative phenotypes, the effect size was determined as the difference between the average phenotype values of carriers and non-carriers. For binary phenotypes, the effect size was the beta coefficient for carrier status from the logistic regression.

#### Common variant analyses

##### Common variant GWAS

We performed genome-wide association analysis on the selected traits using imputed genotypes from the UK Biobank. For each trait we included 13.8 million variants that passed quality control standards (*79*), and further restricted to variants with AF > 0.01. We used self-reported ancestry to exclude non-European participants to reduce confounding due to population structure. Individuals who were closely related to other participants in the UK Biobank cohort were excluded. Each variant was tested for association with the trait under an additive model, using the dosage of the minor allele. Analysis of quantitative traits used phenotype values which had been adjusted for age, sex, and usage of relevant medications, and were inverse rank normalized prior to analysis. In contrast, analysis of binary traits used raw indicator variables. Binary and quantitative traits were both tested with a linear model, and included age, sex, and the first ten genetic principal components in the regressions. The covariates were standardized to mean=0 and standard deviation=1 before regression. We used a P-value threshold of 5×10^-8^ to define genome-wide significance.

##### Fine-mapping of GWAS results

Common variant GWAS results were fine-mapped to target genes which could be responsible for the associations. We used the results from our GWAS association testing in the UK Biobank for most traits, aside for some binary traits with insufficient cases in the UK Biobank and Spherical Equivalent (Nearsightedness), where we used the GWAS Catalog (*84*) as the next best source of association statistics for the trait (Table S1).

To associate significant GWAS variants with protein-coding genes, we first sorted all tested variants that are at least nominally significant at 5×10^-8^ by their P-value. For each variant starting from the most significant, we collected all variants within ±500 kb and performed forward selection stepwise regression against the phenotype values to define a set of independent GWAS signals in the locus. The stopping criterion for the stepwise procedure was when the significance of the remaining variants dropped below 0.05 after correction for multiple testing given the effective number of variants in the locus.

This procedure yielded a set of independent variants that are associated with the phenotype with some potential artifacts at locus boundaries, which may not be truly independent. The initial set of variants was filtered for these artifacts via a second stepwise regression with the same parameters. Next, all remaining variants were grouped into sets of correlated variants based on their pairwise linkage disequilibrium and the most significant GWAS variant in each set was labeled as the index variant for the locus. Two variants were grouped together if the significance of the Pearson correlation between their genotype values in the UK Biobank GWAS cohort was below 10^-6^. Next, each independent variant (both index and secondary variants) was associated with one or more protein-coding genes. To do so, first we considered all common variants in the UK Biobank that were in LD at r^2^ > 0.5 with each independent GWAS variant. In addition, we required that the LD partners of the GWAS variant have INFO scores > 0.4, allele frequency > 0.01, are in Hardy-Weinberg equilibrium (P-value > 1×10^-11^), have at least 50 individuals with imputed genotype calls for each allele within 0.1 of the corresponding hard call (0, 1 or 2), and have P-values that were more than 80% of the index variant P-value when -log_10_-scaled. LD statistics were computed in the UK Biobank GWAS cohort (unrelated white British only) on all 90 million imputed variants. For each LD partner, including the GWAS variant, we determined all genes for which

1) any of the variants in LD is a missense or LoF variant or has SpliceAI score > 0.2 (coding variant), or
2) if no coding variants were found, any of the variants in LD is a known eQTL from CAVIAR (*85*, *86*) with consistent directions of effect across all tissues in GTEx (non-coding variant) or
3) transcription start sites were within the window of 50kb upstream and 50kb downstream of the leftmost and rightmost variant in LD (non-coding variant);
4) if no genes were found according to any of the above rules, we fine-mapped to the gene with the nearest transcription start site to the GWAS variant (non-coding variant)

To fine-map variants from the GWAS catalog, we applied the same procedure except that 1) we did not perform stepwise pruning to consider secondary independent signals in the same locus, and 2) we did not require that variants in LD with the index variant had P-values > 80% of the index variant P-value after -log_10_-scaling since the information needed for these two steps was not available.

Independent GWAS variants with more than 10 associated genes were discarded from further analysis.

#### Rare variant vs common variant analyses

##### Gene-phenotype pairs with significant associations

To determine the number of significant findings at FDR of 0.05, we computed Benjamini-Hochberg FDR based on the P-values from each type of burden test (LoF, missense with and without prioritization, LoF and missense with and without prioritization, and synonymous variants) for all tested gene-phenotype pairs. The FDR correction accounts for the number of tested phenotypes for each type of burden test.

For all gene-phenotype pairs tested in the burden tests for rare variants, we also computed the Spearman correlation between the average phenotype value among all carriers for each missense variant and its PrimateAI-3D score, and the corresponding P-value. To determine the set of 110 gene-phenotype pairs with significant correlations (Table S9), we computed Benjamini-Hochberg FDR on all genes-phenotype pairs that had at least 10 missense variants with associated phenotype values. The FDR correction accounts for the number of tested phenotypes.

##### Comparison to Backman et al.

Backman et al. used burden tests implemented by REGENIE (*18*) with different allele frequency masks ranging from singleton variants to 1% allele frequency and reported results at 5% Bonferroni corrected P-value threshold of 2.18×10^-11^. To compare our findings to Backman et al. we tried to match the variants used in both pipelines and our significance thresholds as closely as possible. For this reason, we compared our findings for deleterious variants (LoFs and missense variants ranked by PrimateAI-3D) to all significant gene-phenotype associations from Backman et al. for rare missense variants filtered by five pathogenicity predictors (SIFT, PolyPhen2 HDIV, PolyPhen2 HVAR, LTR and MutationTaster) + LoFs variants at all allele frequency thresholds up to 0.1% (corresponding to masks 'M3.singleton', 'M3.0001', 'M3.001' and 'M3.01' from Backman et al.) across all tested ethnicities. We subset the findings from Bachman et al. to 74 phenotypes that were analyzed in both studies (Table S6). Since rare variant testing in Backman et al. was performed and reported on each ethnicity separately, we required that the findings from this study pass the reported P-value significance threshold of 2.18×10^-11^ in at least one of the tested ethnicities. To calculate a matching P-value threshold for our pipeline, we computed a Bonferroni corrected P-value threshold for performing the same number of tests per gene minus the four masks that we considered for missense + LoFs from Backman et al. This P-value threshold is very conservative with respect to our pipeline since it assumes that we would also perform single variant tests, and test five masks for LoF variants and one mask (for 1% allele frequency) for missense + LoF variants for each gene as was done in Backman et al. The estimate does not penalize our pipeline for testing four masks for allele frequencies up to 0.1% for missense + LoF variants, since our pipeline calibrates for this automatically via permutation tests (see *Rare variant burden test* section above). The resulted Bonferroni corrected P-value threshold for the gene-phenotype associations from our pipeline was 2.35×10^-11^, which was very close to the threshold from Backman et al. We further required that the gene-phenotype associations for both pipelines have HGNC gene symbols reported in GENCODE v39.

To compute the percentage of OMIM genes found by each method, we selected the most significant UK Biobank phenotype for each significant gene-phenotype association reported by either our pipeline or by Backman et al. We intersected these genes with the OMIM database to derive triplets of gene, UK Biobank and OMIM phenotypes. We noticed that in some cases, the UK Biobank phenotypes were not clinically related to the OMIM phenotypes linked to the same gene. For this purpose, we curated manually which UK Biobank phenotypes were a good match for the OMIM phenotypes linked to the same gene (Table S7). The reported percentages in Fig. 1C are based only on genes with matching UK Biobank and OMIM phenotypes.

##### Evaluation of missense variant prioritization methods

We derived a high confidence set of gene-phenotype pairs (Table S8) by selecting genes that were significant on the burden test for both LoFs and for total missense variants without prioritization at a nominal P-value threshold of 10^-12^. If a gene was associated with multiple phenotypes, the phenotype with the most significant P-value by the burden test for total missense variants was kept and the other phenotypes were discarded.

Multiple missense classifiers were considered for pathogenicity prediction in the burden tests, including BayesDel (*67*), CADD (*68*), ClinPred (*69*), DEOGEN2 (*69*), EVE* (*61*), FATHMM-XF (*70*), M-CAP (*71*), MetaLR (*72*), MetaSVM (*72*), MutationAssessor (*73*), Polyphen-2 (*74*), PrimateAI-3D (*12*), PROVEAN (*75*), REVEL (*76*), SIFT (*77*), and VEST4 (*78*). Pathogenicity scores for the EVE-style variational autoencoder (EVE*) were generated by re-implementing the method (*61*). The scores for all other methods were downloaded from dbNSFP v4 (*87*, *88*). For the evaluations in Figs. 1D, S2 and 3D we required that the variants are scored by all methods and that each gene has at least 10 such variants.

In Fig. 1D, to estimate an upper bound of the performance for each gene, conceptually, we treated carriers of the same missense variant as replicate observations of the phenotype. For this purpose, we used only variants with at least four carriers, which reduced the total number of variants for each gene since we had to discard variants with fewer carriers. In total, 62 genes were used, for which there were at least 10 such variants that were scored by all methods. For each gene, we created 1,000 pairs of test and training sets by randomly selecting one of the carriers for each variant to be in the test set and the rest of the carriers for that variant to be in the training set. Then, we measured how well each training set predicts the corresponding test set and compared this to the prediction accuracy of each method for missense variant prioritization. For each variant we computed the average phenotype value among all carriers in the training set and correlated these values using Spearman correlation with the phenotype values in the training set. In the same way, we also correlated the pathogenicity scores from each method with the phenotype values in the test set. For each score, we reversed the sign of the correlation if rare LoF variants were predicted to decrease the phenotype in our burden test, so that positive correlations indicate agreement between the direction of effects of LoFs and the predicted pathogenicity of missense variants, whereas negative correlations indicate disagreement. The mean correlation among all 1,000 splits was computed for each gene and method and used as input to Fig. 1D. In Fig. S3 we estimated the fraction of variance explained by computing the square of the mean Spearman correlation for each gene. Furthermore, we compared all scoring methods to two lower bound benchmarks: 1) a uniformly random score in the range between 0 and 1 for the same set of missense variants; and 2) variance explained by using phenotype values of carriers of the same synonymous variant for synonymous variants with at least four carriers in the same gene. Spearman correlations were compared between PrimateAI-3D and other methods by a Wilcoxon signed-rank test.

In Fig. 3D, European variants were defined as missense variants with MAF < 0.1% in Europeans and allele count of 0 in non-Europeans in the UK Biobank. Conversely, non-European variants were defined as MAF < 0.1% in non-Europeans and allele count of 0 in Europeans. Ultra-rare non-European variants were defined as missense variants with allele count of 1 in non-Europeans and allele count of 0 in Europeans in the UK Biobank. Ultra-rare European variants were defined as missense variants with allele count of up to 24 in Europeans and allele count of 0 in non-Europeans. The allele count threshold for ultra-rare European variants of 24 was set to be equal to the ratio between European and non-European individuals with exome data in the UK Biobank, which was 24.6. Additionally, ultra-rare variants were required to be absent from TOPMed. We assessed correlations between pathogenicity score and phenotype in carriers. Each gene was required to have at least 10 variants with at least one carrier with phenotype values and pathogenicity scores across all methods in all tested group (rare and ultra-rare in Europeans, rare and ultra-rare in non-Europeans). We excluded EVE* from this comparison, because this method caused too many genes to be discarded due to missing scores for ultra-rare non-European variants. The final set of genes in Fig. 3D consisted of 49 genes. Performance of PrimateAI-3D scores between different sets of variants was compared with two-sided t-tests for related samples between the Spearman correlations across all genes from the corresponding pairs of sets of variants.

##### Rare variants enrichment in GWAS genes

In Fig. 4F, for each pair of phenotypes, we tested the statistical significance of the enrichment of rare variants affecting the first phenotype in GWAS genes associated with the second phenotype. We first sorted all protein-coding genes in the genome by the P-value of the rare variants burden test for the first phenotype and assigned a rank to each gene, G_i_, where 1 <= G_i_ < =N (total number of genes) and where G_i_ = 1 for the most significant gene and G_i_ = N for the least significant gene. Next, we checked whether any subset of the fine-mapped GWAS genes for the second phenotype ranks significantly higher than expected by chance. For this purpose, we sorted all GWAS genes by their ranks from the burden test and incrementally used a series of hypergeometric tests to compute the likelihood of observing by chance the same number of genes that rank at least as highly as in the observed data. Formally, let K be the total number of GWAS genes for the second phenotype and [G^GWAS^_1_, G^GWAS^_2_ , …, G^GWAS^_K_], be the sorted list of ranks of the GWAS genes according to the burden test for the first phenotype. We performed K successive hypergeometric tests to produce an array of P-values [P_1_, P_2_, …, P_K_], where the *i*-th hypergeometric test is testing the probability to observe by chance *i* genes out of K, where the *i*-th gene ranks at most G^GWAS^_i_ in a set of N total genes. To account for multiple testing (K hypergeometric tests), we applied Benjamini-Hochberg False Discovery Rate correction on the above array of P-values and picked the most significant one as the significance of the enrichment of rare variants affecting the first phenotype in GWAS genes for the second phenotype. We recorded the number of GWAS genes in the subset that produced the best FDR-corrected P-value and also the percentile of the subset in the ranking of all genes in the genome. The magnitude of the effect (corresponding to the size of the circles in Fig. 4F) was computed as the observed overlap divided by the expected overlap for the most significant overlap determined by the above procedure. For visualization purposes, the size of the circles in the figure was set to 1 + log_10_(magnitude) and rescaled by the maximum across the whole matrix.

##### Variant effect size as function of allele frequency

To show the relationship between variant effect size and allele frequency, we selected all genes among those detected by our GWAS fine-mapping pipeline which were also significantly enriched for deleterious rare variants based on the burden test at FDR of 5%. If a gene was associated with multiple phenotypes, the gene-phenotype pair with the most significant GWAS P-value was used and the rest of the phenotypes were discarded. Furthermore, we excluded gene-phenotype pairs for which our rare variants burden tests for deleterious variants and for LoF variants had opposing directions of effects.

Directly plotting effect sizes as function of allele frequency across multiple genes and phenotypes can introduce substantial noise, because, depending on the exact relationship between each specific gene and phenotype, breaking the function of some genes may cause phenotype values to decrease whereas breaking other genes may cause the opposite. To alleviate this problem, we considered the direction of effect of LoF variants in our rare variants burden test and reversed the sign of the effects of all variants in genes where rare LoFs collectively caused a decrease in the phenotype. For common GWAS variants, we considered only the most significant variant for each gene and computed the effect size with respect to its minor allele. Similarly to exome variants, we reversed the sign of common GWAS variants in genes where rare LoFs were predicted to decrease the phenotype by our burden tests.

After the above adjustments, variants were grouped by log_10_ minor allele frequencies into 11 equally spaced bins and the average effect size across all variants in each bin was computed. To model the relationship between effect size and minor allele frequency for each variant category (LoF, deleterious missense, etc) we used a similar approach to the “alpha model” from (*27*, *28*), which relates effect size, β, to minor allele frequency, *p*, via:

$$E(\beta^{2}|p) =\sigma^{2} \left[ 2p(1-p) \right]^{\alpha}$$

where $\sigma$ and $\alpha$ are parameters to be estimated from the data. In our case, we modelled the raw effect size, β, as

$$E(\beta|p) \sim\sigma\sqrt{\left[ 2p(1-p) \right]^{\alpha}}$$

A generalized linear model with Gaussian distribution and log link function was fitted to estimate the parameters for each variant category.

Deleterious and non-deleterious missense variants were defined as missense variants with PrimateAI-3D scores in the top 80% and bottom 20%, respectively, within each gene. Cryptic splicing variants were defined as variants with SpliceAI score > 0.2 that are outside of the canonical splice junctions.

##### Fraction of GWAS loci with rare variants signal

In Fig. 4G, we selected GWAS loci where the index variant was associated unambiguously with a single protein-coding gene either via coding or non-coding variants in LD with the index variant. If a gene was associated with multiple phenotypes, we kept the gene-phenotype pair with the most significant GWAS P-value and discarded the rest. We binned GWAS loci by P-value and computed the fraction of genes for which our burden test for deleterious variants produced a nominal P-value ≤ 0.05. To define “High confidence” GWAS variants, we selected genes-phenotype pairs with GWAS P-values < 10^-100^, for which the index variant was in very high linkage disequilibrium (r^2^ ≥ 0.9) with the coding variants identified during fine-mapping. We also created a category of genes which excluded genes with short length (coding sequence <2 kb) and genes under strong evolutionary selection (pLI score > 0.99). Spearman correlation was computed between burden test P-values and GWAS P-values, pLI scores and coding sequence length for genes implicated by both GWAS and rare variant burden tests.

#### Polygenic risk scores

##### Common variant polygenic risk scores

Polygenic risk scores (PRS) were constructed from common and rare variant tests at different P-value thresholds. We reduced the 90 traits by excluding six underpowered binary traits, along with twelve of the more redundant traits. We constructed the common variant PRS from independent significant variants for each trait. For this we ranked all variants by their GWAS P-value, selected the variant with the strongest P-value, excluded variants within 1 Mb of that index variant, and repeated selecting the strongest remaining variant and excluding around that until none achieved the P-value threshold. We also excluded variants in strong LD to index variants (UK Biobank Europeans r^2^ > 0.1), if they were within 10 Mb of any previously chosen index variant.

We permitted multiple independent signals within a locus by first including the most significant variants via a forward-stepwise regression. This involved selecting all common variants with GWAS P-values within 1 Mb of the index variant, regressing each variant against the trait, selecting the most significant variant, then substituting the trait with the most significant variant’s residuals. This was repeated until the log-transformed P-value of the most significant variant was less than 25% of the index variant log_10_-scaled P-value, or three variants had been selected, whichever came first.

The variants in a locus identified by the stepwise regression were further pruned by performing a multiple linear regression with all variants versus the trait, and retaining only variants which remained more significant than our P-value threshold. The regression effect sizes and P-values were stored, and PRS computed, which for an individual was the sum of effect size × dosage from each independent variant.

##### Rare variant polygenic risk scores

We constructed the rare variant PRS from the results of the rare variant burden tests. These were conducted per gene, and each gene had separate thresholds for rarity (allele frequency) and pathogenicity (e.g. PrimateAI-3D) established in the training group. For genes with burden test P-values more significant than the required threshold (typically 1×10^-6^), we identified variants predicted to be sufficiently pathogenic (loss-of-function consequence or SpliceAI > 0.2, or PrimateAI-3D > gene pathogenicity threshold).

We filtered these variants by rarity, requiring gnomAD and TOPMed allele frequencies to be less than or equal to our per-gene threshold. We also required the UK Biobank test group allele frequencies to not significantly exceed the threshold from the training group. This was conducted separately for the different ancestral groups in the UK Biobank, excluding the variant if any ancestry group failed this criteria. The groups checked were African (AFR), East Asian (EAS), European (EUR) and South Asian (SAS) ancestries, from self-determined labels. The AFR, EAS and SAS groups included all possible individuals for each group, as none of these individuals were included during training, whereas the EUR testing group excluded the training individuals. For each ancestry group we compared the number of alternative and reference alleles between the testing group and the training group via a one-tailed Fisher’s exact test, and excluded variants where any ancestry group had a significantly higher alternative allele frequency compared to the training group with P < 0.05.

We modelled the effects of predicted pathogenicity and allele frequency phenotype in the training group to better predict individual’s phenotypes. Using the sufficiently rare and predicted deleterious variants according to our per-gene thresholds, we regressed predicted pathogenicity scores and log_10_ allele frequencies against phenotype values. For individuals with multiple rare deleterious variants per gene, we selected the variant with the highest predicted pathogenicity score. This gave a model which could be used to predict a per-gene phenotype, given a variant’s predicted pathogenicity and allele frequency.

We predicted these values for individuals in the training group who were heterozygous or homozygous for the alternate allele for one of the rare and deleterious variants. These values were then reweighted according to our confidence in the gene. We reasoned that genes with less certain effect sizes were less reliable. We constructed 95% confidence intervals around each gene’s effect size, using the effect size and standard error of the effect size from the rare variant burden test. If the 95% CI included 0, the assigned weight was 0, otherwise the weight assigned to a gene was the ratio between the CI bound nearest zero, and the original effect size. The rare variant PRS per individual was the sum of the reweighted values across all significant genes.

##### Polygenic risk score assessment

We illustrated the PRSs constructed from rare variants for total cholesterol levels. We selected the significant genes used for the rare variant PRS. We identified the pathways for cholesterol metabolism, and their pertinent cells and tissues. A diagram of these cholesterol metabolic pathways was created with [BioRender.com](http://www.biorender.com), and annotated with the genes significant in the rare variant burden test, along with their effect direction and size.

We evaluated PRS predictions for the quantitative traits in several ways. For two traits, HbA1c and LDL cholesterol, clinical diagnostic criteria were available which we used to classify cases from controls for type 2 diabetes and dyslipidemia, respectively. For each trait, we ranked individuals by the respective PRS (for common, rare and unified variant models), and counted the number which could be identified at different odds ratios. Due to individuals with extremely low PRS predictions having very low odds of disease the distribution of odds ratios peaked twice, at either end of the PRS spectrum, firstly near individuals with the lowest PRS predictions, and secondly in individuals with the highest PRS predictions. Following the initial peak, the odds ratio declined slowly to a minimum before rising again after the cases in the upper group became sufficiently enriched. Consequently, we used the local minimum between the two end peaks as our baseline for selecting at-risk individuals. We tested whether the unified variant PRS model identified significantly more individuals than the common variant PRS model alone by Fisher’s exact test.

We also assessed how well the different missense pathogenicity prediction models performed when used for rare variant burden tests and subsequent construction of rare variant PRS predictions. For each pathogenicity prediction we calculated the correlations with the original phenotype in a subset of individuals held out from training. We plotted the mean correlation across the quantitative traits, along with the mean P-value after log_10_ transformation. Subsequent assessments use results for PrimateAI-3D only, as PrimateAI-3D had the highest mean correlation with original phenotype. We compared the correlation found with the rare variant PRS models to that found with common variants, and a unified model which summed common and rare variant PRS predictions together. The third evaluation tested enrichment of PRS outliers across a range of z-score phenotype outlier cutoffs. For each of these cutoffs we identified individuals above the phenotypic cutoff, and determined the proportion of those with an outlier PRS score. We assessed this at two PRS thresholds: 99% and 99.9%. We averaged proportions across phenotypes, and scaled values relative to the phenotypic z-score=0 cutoff, i.e. full cohort proportion. We tested for differences in enrichment between common and rare variant PRS at the most extreme phenotype threshold by two-sample t-test. Lastly, we compared enrichment of PRS outlier individuals within phenotypic outliers by Fisher’s exact test. We tested enrichment at three thresholds for defining outliers: 10%, 1% and 0.1% (for example defining the top 10% and bottom 10% as outliers). We counted the fraction of phenotypes with a P-value below a Bonferroni corrected threshold of 0.05, and tested for a difference between rare and common variant PRS models by Fisher’s Exact test.

We evaluated the performance of the PRS models among individuals with non-European ancestry. Using self-declared ancestry, we grouped individuals into one of four ancestries – African, East Asian, European, or South Asian. None of the individuals with non-European ancestry were included during training, and so were all included for testing. We constructed common variant and rare variant PRS predictions for the test individuals. For modelling per-variant effects in the rare variant PRS, we used Lasso regularization via a model which included all significant genes, to better account for overfitting. The Lasso model used five-fold cross-validation to estimate a lambda value from the optimal lambda plus one standard error. We counted the number of variants per individual when constructing each rare variant PRS, to verify this matched between ancestries. We calculated the Pearson correlation between PRS predictions and phenotype values. We scaled the non-European correlation relative to the correlation observed in individuals with European ancestry, and tested whether the correlations across all phenotypes was significantly below 100% (relative to Europeans) using a one-sample t-test. In addition to evaluating relative correlation, we also evaluated the ability to discriminate phenotypes at the rare variant PRS extremes. We identified individuals in the bottom 0.5% and top 0.5% of the PRS, and calculated the difference between the mean phenotype values for each group. We compared this distance between individuals with European vs non-European ancestries.

We tested the reproducibility of the rare variant PRS in an independent cohort - MGB, containing 20,708 individuals with exome-based genotypes, and who were phenotyped for 17 quantitative traits also present in the UK Biobank. We trained rare variant PRSs using all of the UK Biobank unrelated European individuals with exome data, and identified rare variants in relevant genes in the MGB cohort, given our PrimateAI-3D and allele count thresholds per gene. We created risk scores for the 17 traits, and evaluated performance via correlation with the original phenotype, as well as assessing enrichment of PRS outliers in phenotype outliers for each trait using a threshold of 1% to define outliers. The same evaluations were also done for the UK Biobank, but with 50% train, 50% test groups.

**Forecasting analysis**

We randomly down-sampled the UK Biobank exome cohort. We ran burden tests with PrimateAI-3D and GWASes on each subset of individuals for the 79 quantitative phenotypes in our study. We identified rare variant-associated genes at FDR < 5% (Fig. 4A) and independent GWAS significant loci at P < 5×10^-8^ (Fig. S12), as previously described. In Fig. 4B, for each variant type (LoFs, deleterious missenses and the union of LoFs and deleterious missenses) in all significant genes on the burden test, we counted the number of variants that passed the optimal allele count and PrimateAI-3D thresholds from the burden test and divided the count by the number of individuals in the down-sampled cohort. To avoid double-counting, variants in genes associated with multiple phenotypes were only included once. In Fig. 4C, we computed the average absolute effect size of newly discovered genes at each cohort size. In Fig. 4D, we trained rare variant polygenic scores and plotted the median correlation with phenotypes at each cohort size. In Fig. 4E, we determined proper subsets of genes that were significant on our burden test (FDR < 5%) at incrementing cohort sizes (i.e. genes significant at larger cohort sizes, but not significant at smaller cohort sizes were discarded from this analysis). The areas of the circles for rare variant genes were drawn proportional to the number of significant rare variant-associated genes at each cohort size. The area of the circle for GWAS loci was drawn proportional to the total number of index variants when using the complete UK Biobank cohort across all phenotypes. The areas of the overlaps between rare variant genes and GWAS loci were drawn proportional to the number of significant rare variant-associated genes at each cohort size whose transcription start sites were within 1 megabase of any of the local independent GWAS variants for the same phenotype when using the complete UK Biobank cohort.

### Supplementary figures

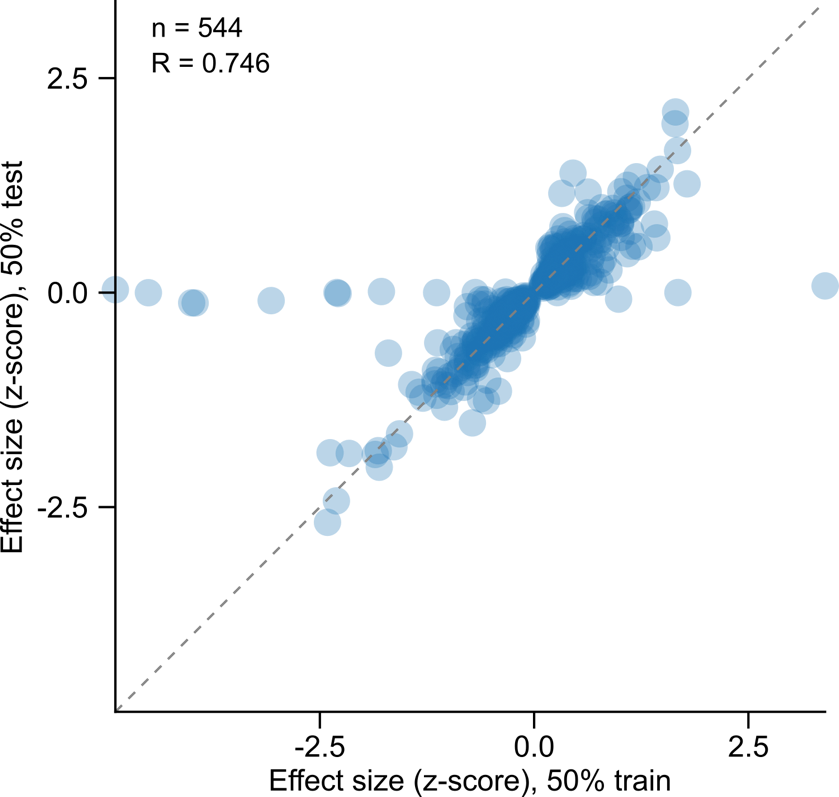

**Fig. S1. Replication of effect sizes in a held out subset**. Rare variant tests were performed using 50% of the cohort as a training set, and the other 50% as a testing set. Effect sizes are compared from the training set versus the test set for genes with a burden test P-value < 10^-7^ in the training set.

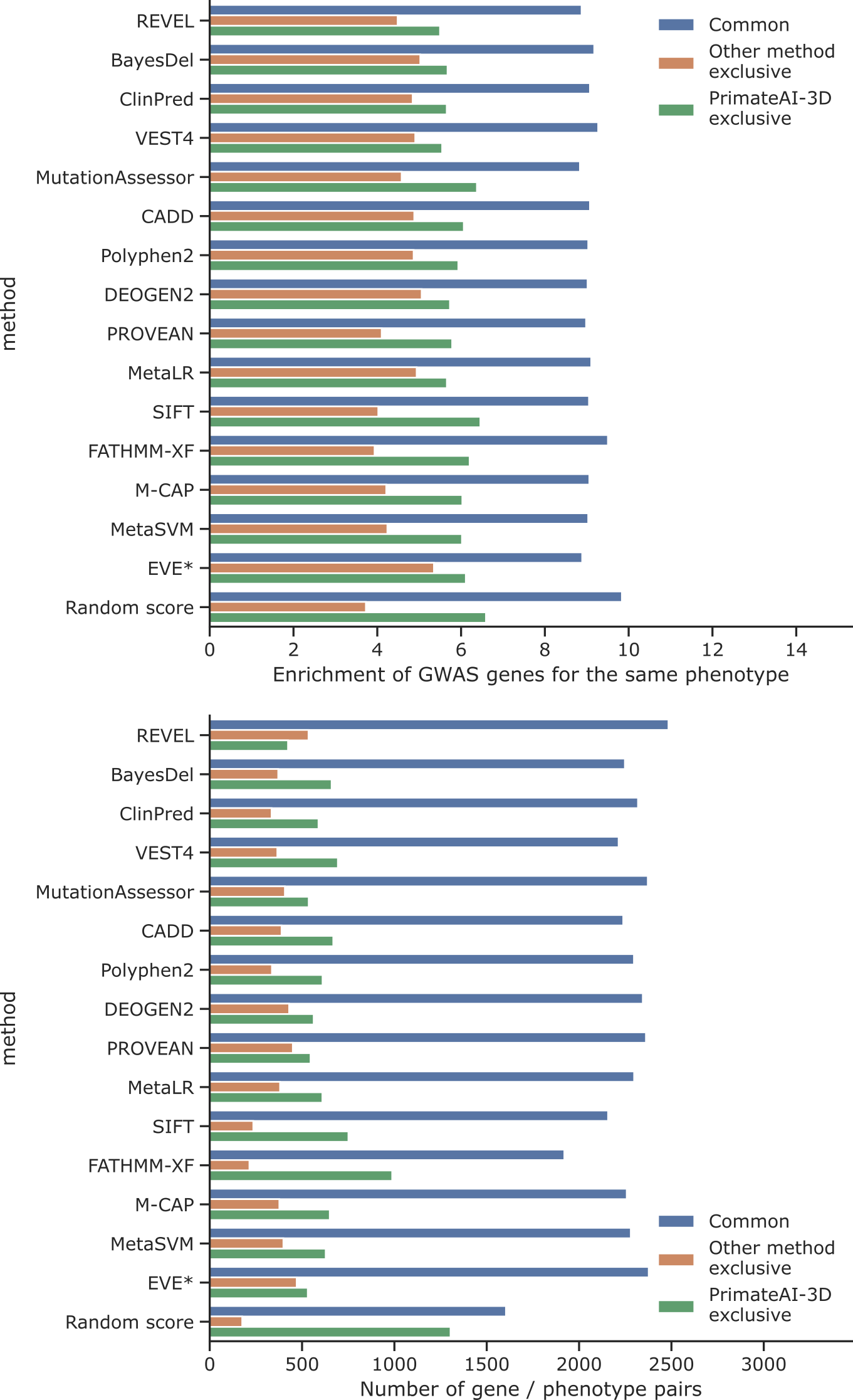

**Fig. S2.** **GWAS enrichments and number of findings for different pathogenicity classifiers**. For each method, we partitioned the gene/phenotype pairs into three groups: 1) pairs detected by both PrimateAI-3D and the other method (e.g. REVEL, BayesDel etc); 2) pairs detected exclusively by the other method and not by PrimateAI-3D; and 3) pairs detected exclusively by PrimateAI-3D and not by the other method. The enrichment of GWAS genes for the same trait (upper plot) and the number of findings (lower plot) are shown for each group. Gene-phenotype pairs exclusively detected by PrimateAI-3D show consistently higher enrichments of GWAS genes for the same trait. The results for a random pathogenicity score are shown as negative control.

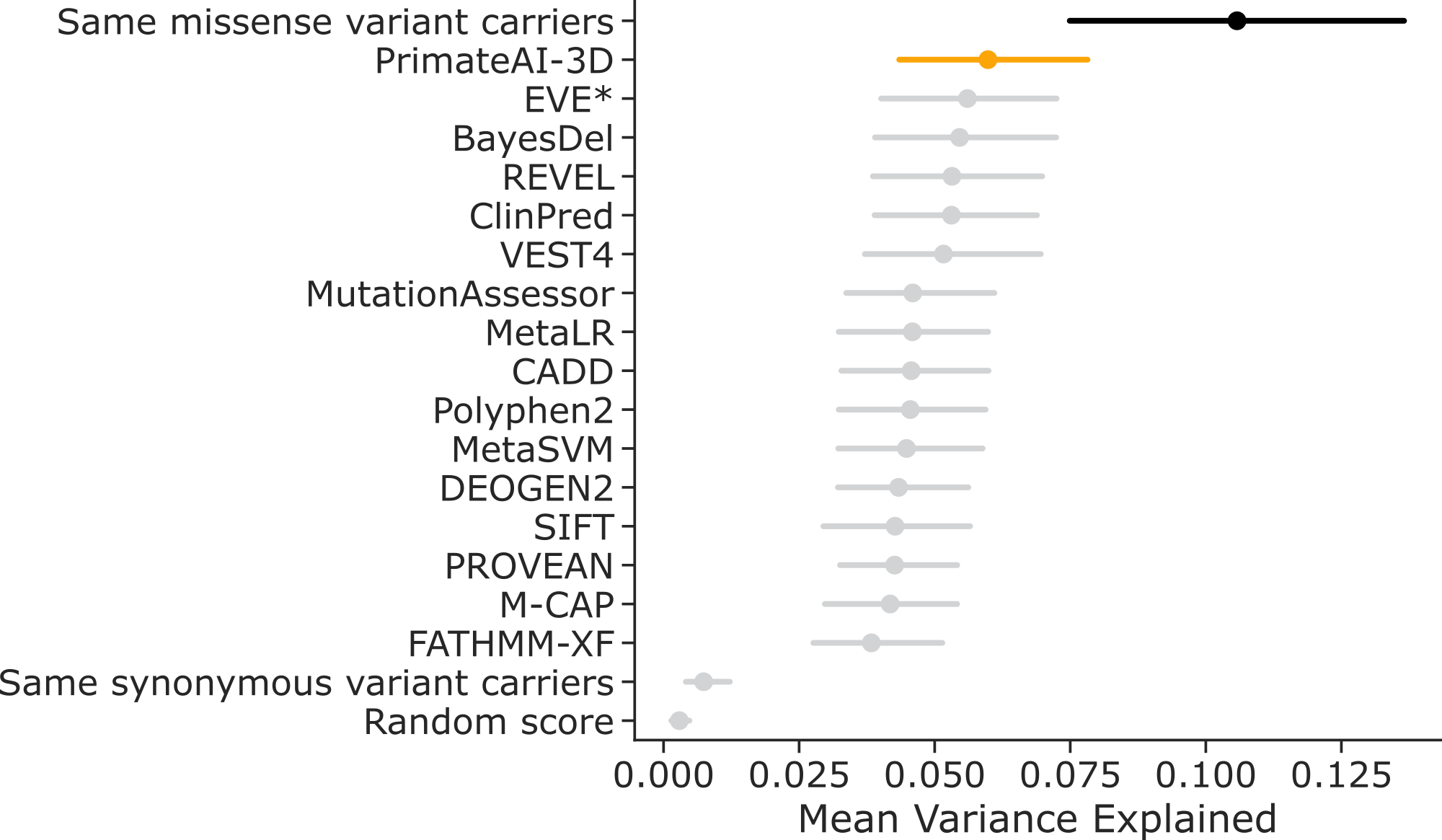

**Fig. S3. Mean fraction of phenotypic variance explained by different pathogenicity scoring methods.** Methods were evaluated on a set of 62 gene-phenotype pairs, which were selected based on their enrichment of rare LoF and missense variants without prioritization (Fig. 1D). The phenotypic variance explained is computed as the squared Spearman correlation between the scores from each scoring method and the phenotypic values of the carriers of the corresponding variants. The methods are compared to a theoretical upper bound, which was computed by using phenotypic values of carriers of the same missense variants, and to two theoretical lower bounds - carriers of the same synonymous variants and random scores. Error bars represent 95% confidence intervals.

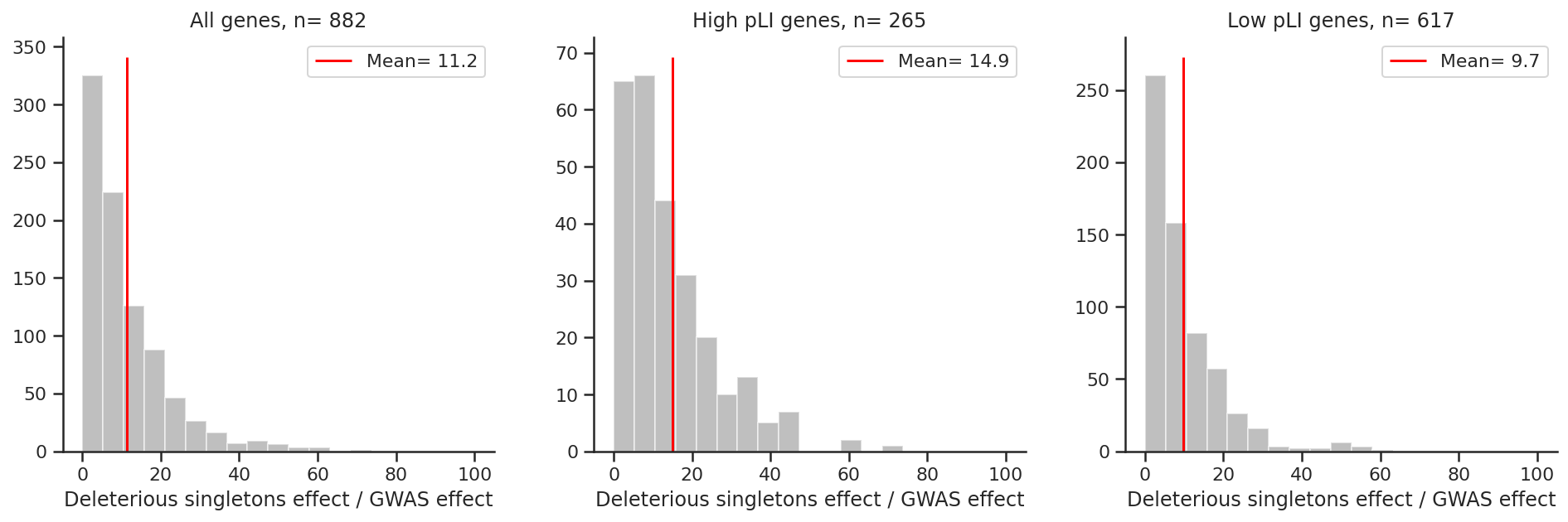

**Fig. S4**. **Effect size ratios between rare and GWAS variants.** Ratios between the effect sizes of the rarest deleterious variants in the UK Biobank and the most significant GWAS variant for genes implied by both GWAS and rare variant analysis. Effects of the rarest deleterious variants in the UK Biobank exome cohort are on average 11.2-fold larger than the corresponding sentinel GWAS variants for the same gene (**left plot**). In genes under strong evolutionary selection (pLI score > 0.5, **middle plot**), this ratio was 14.9-fold compared to 9.7-fold for genes under weak selection (pLI score < 0.5, **left plot**, rank-sum P-value = 6×10^-10^).

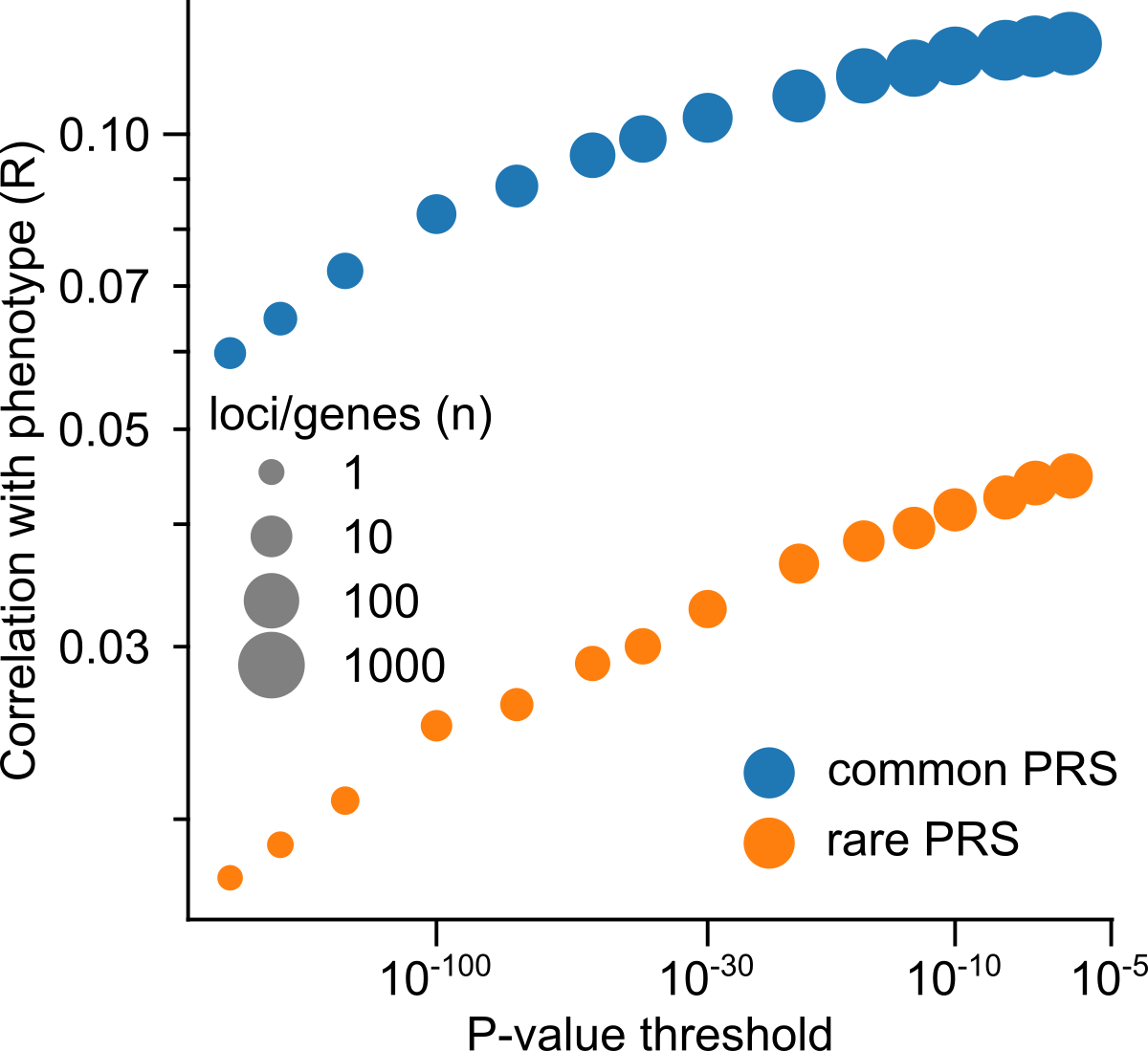

**Fig. S5. Common variant and rare variant PRS subsets.** Median common and rare variant PRS correlations when loci were required to be significant in both common and rare tests. PRSs were trained on 90%, tested on the remainder.

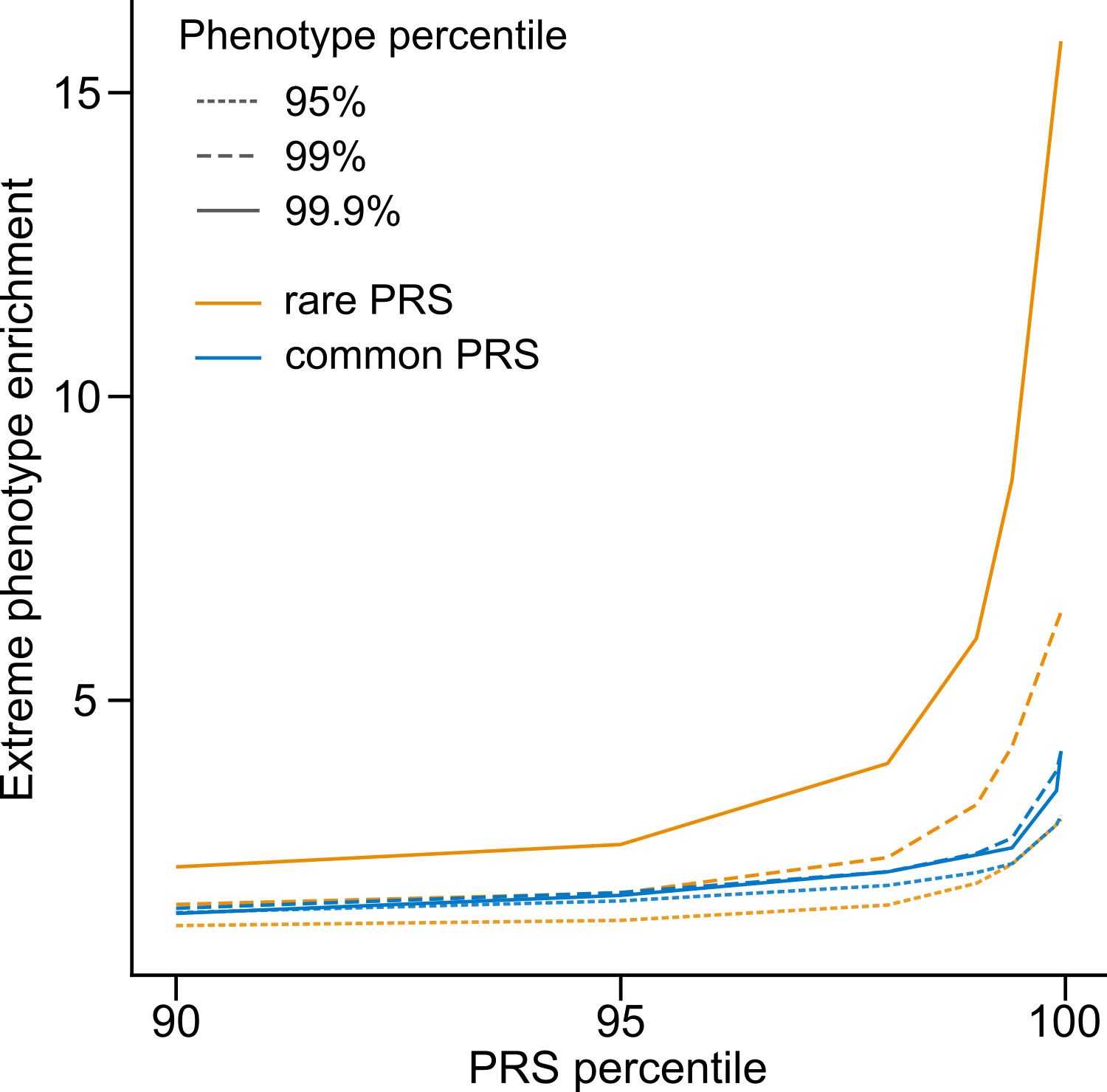

**Fig. S6. Extreme phenotype enrichment for individuals with an extreme PRS.** Stated PRS and phenotype percentiles also include low extremes, for example, the 95% percentiles include individuals in the top 5% and bottom 5%. Enrichment is shown compared to the fraction expected from the percentile.

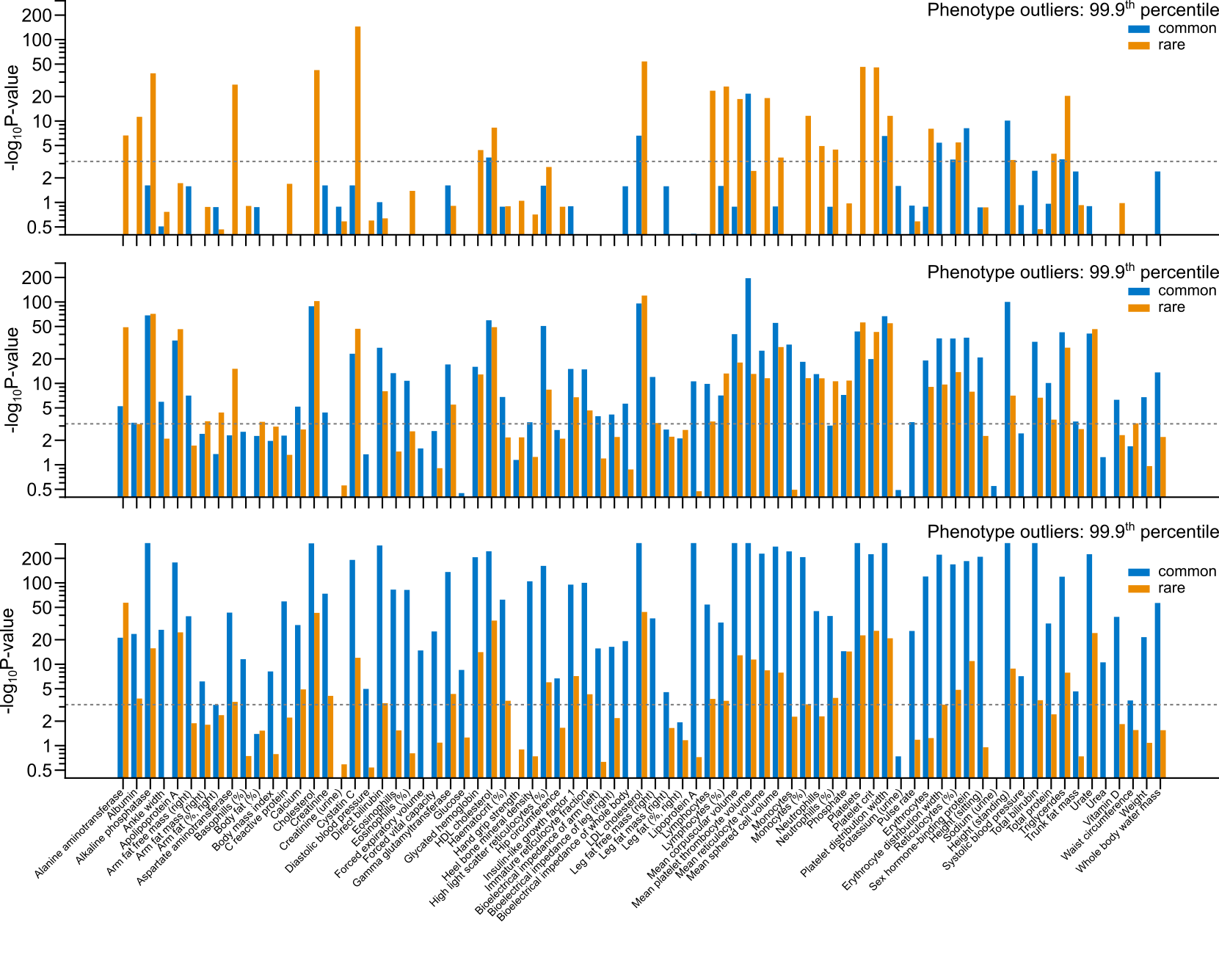

**Fig. S7. PRS outliers by trait for all 78 tested quantitative traits**. Bars indicate statistical significance per trait for the common and rare variant PRSs, across three PRS and phenotype outlier thresholds: 90^th^, 99^th^ and 99.9^th^ percentiles. PRSs were trained on 50%, tested on the remainder.

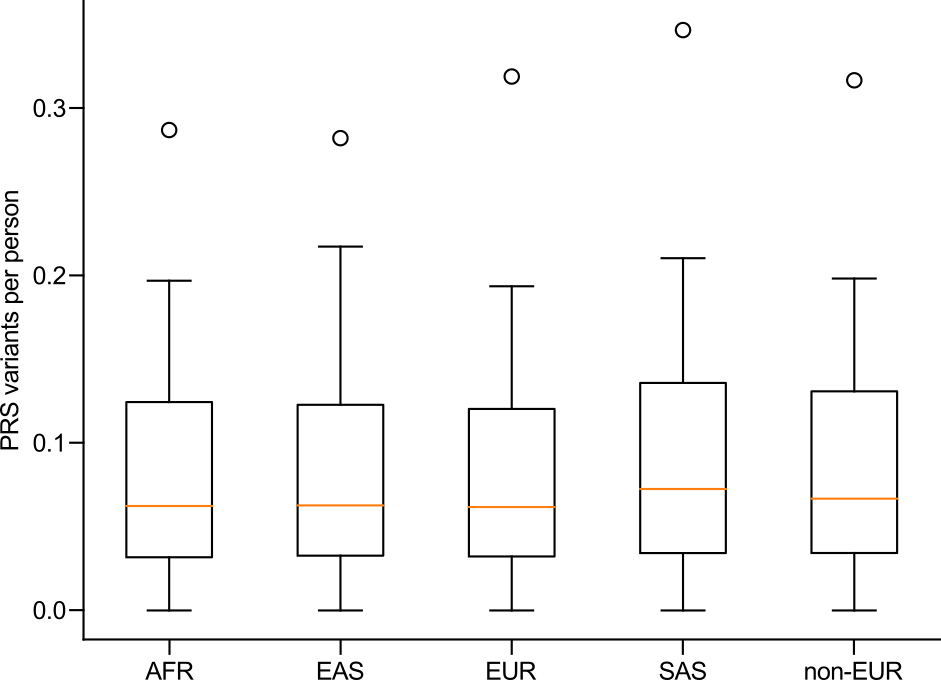

**Fig. S8. Number of variants per person when constructing rare variant PRS, by ancestry.** AFR: African, EAS: East Asian, EUR: European, SAS: South Asian , non-EUR: non-European.

**Fig. S9. Rare variant PRS distributions by ancestry.** Distributions of PRS scores within different ancestry groups from the UK Biobank, for the quantitative traits. AFR: African, EAS: East Asian, EUR: European, SAS: South Asian, non-EUR: non-European.

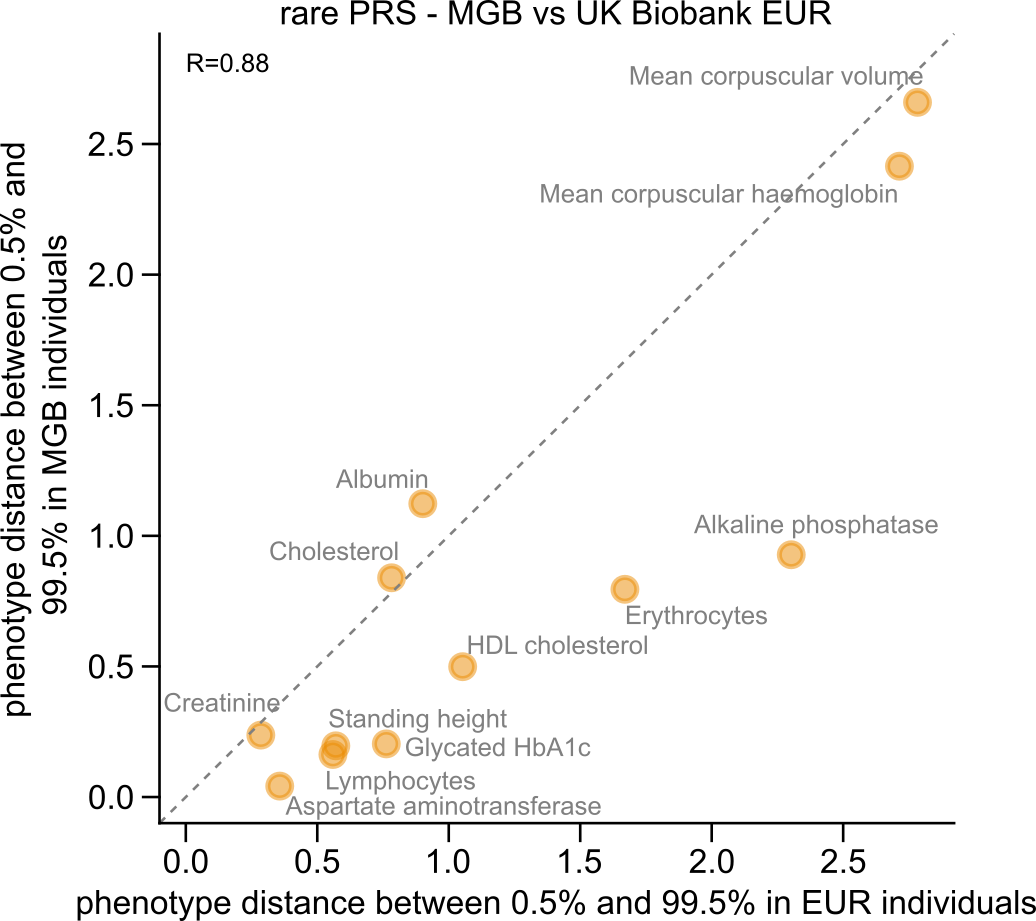

**Fig. S10. Rare variant PRS portability between cohorts.** Mean phenotype distance (z-score) was calculated for individuals with low (<0.5%) and high (>99.5%) rare variant PRS percentiles. Scatterplot compares mean phenotype distance in UK Biobank EUR (x-axis) and MGB (y-axis) individuals for 16 traits. A line of equivalence is shown by the gray diagonal dashed line.

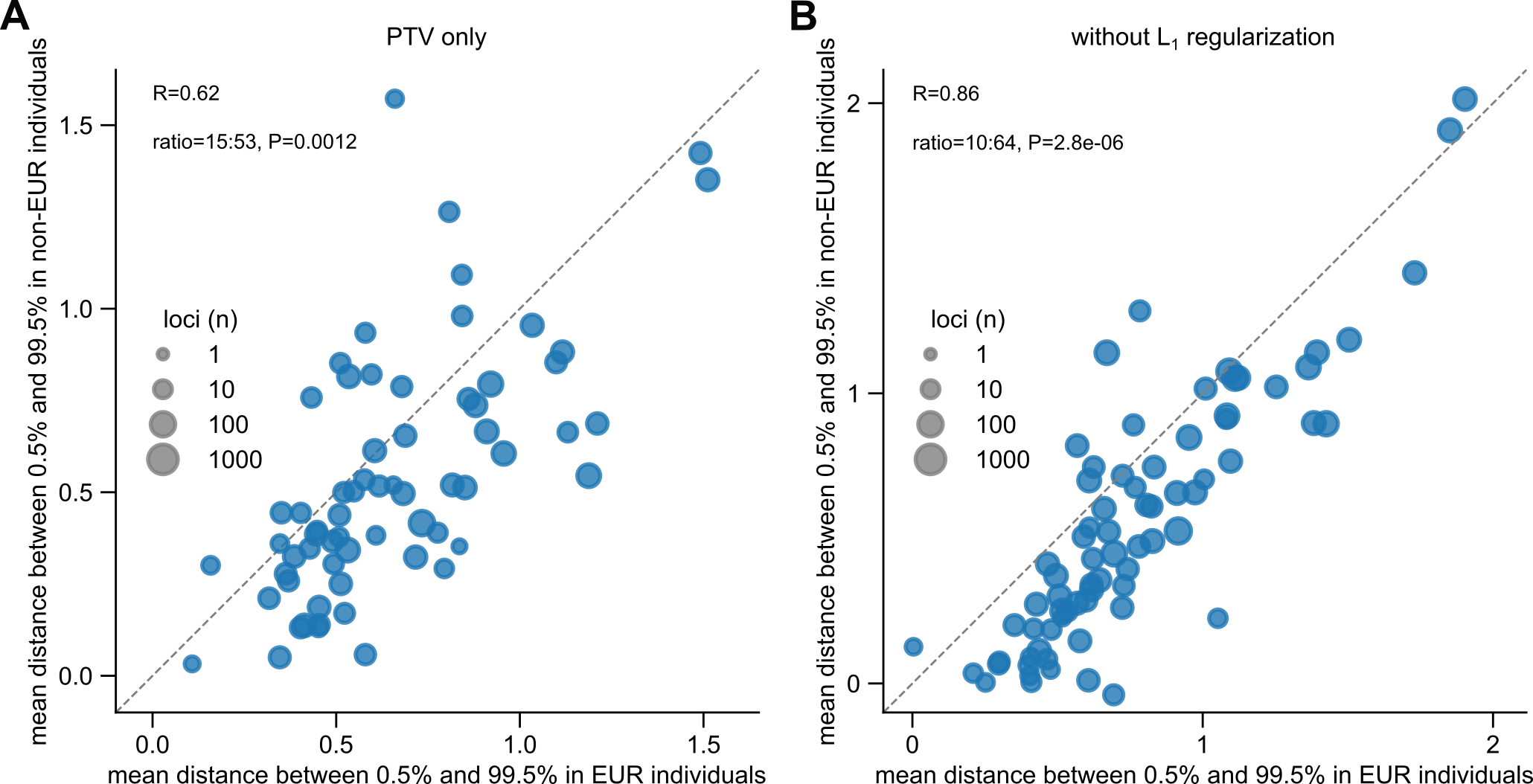

**Fig. S11. Rare variant PRS by ancestry.** (**A**) Comparison of mean z-score phenotype distance between low PRS (<0.5%) and high PRS (>99.5%) groups, but restricted to PTVs only. (**C**) Comparison of mean z-score phenotype distance as in (B) by trait, for both EUR and non-EUR individuals, without using L_1_ regularization. Lines of equivalence are shown by the gray diagonal dashed line.

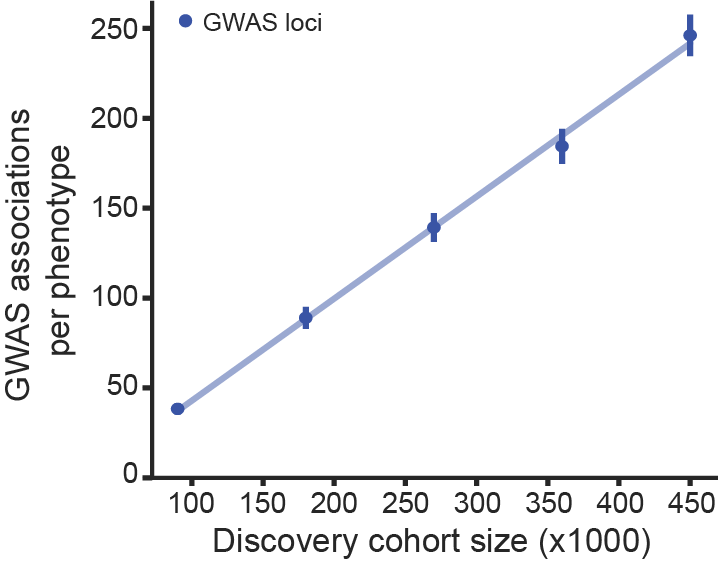

**Fig. S12**. Number of genome-wide significant GWAS loci identified per phenotype as a function of the discovery cohort size in thousands of individuals. Dots and error bars represent mean ± standard error.

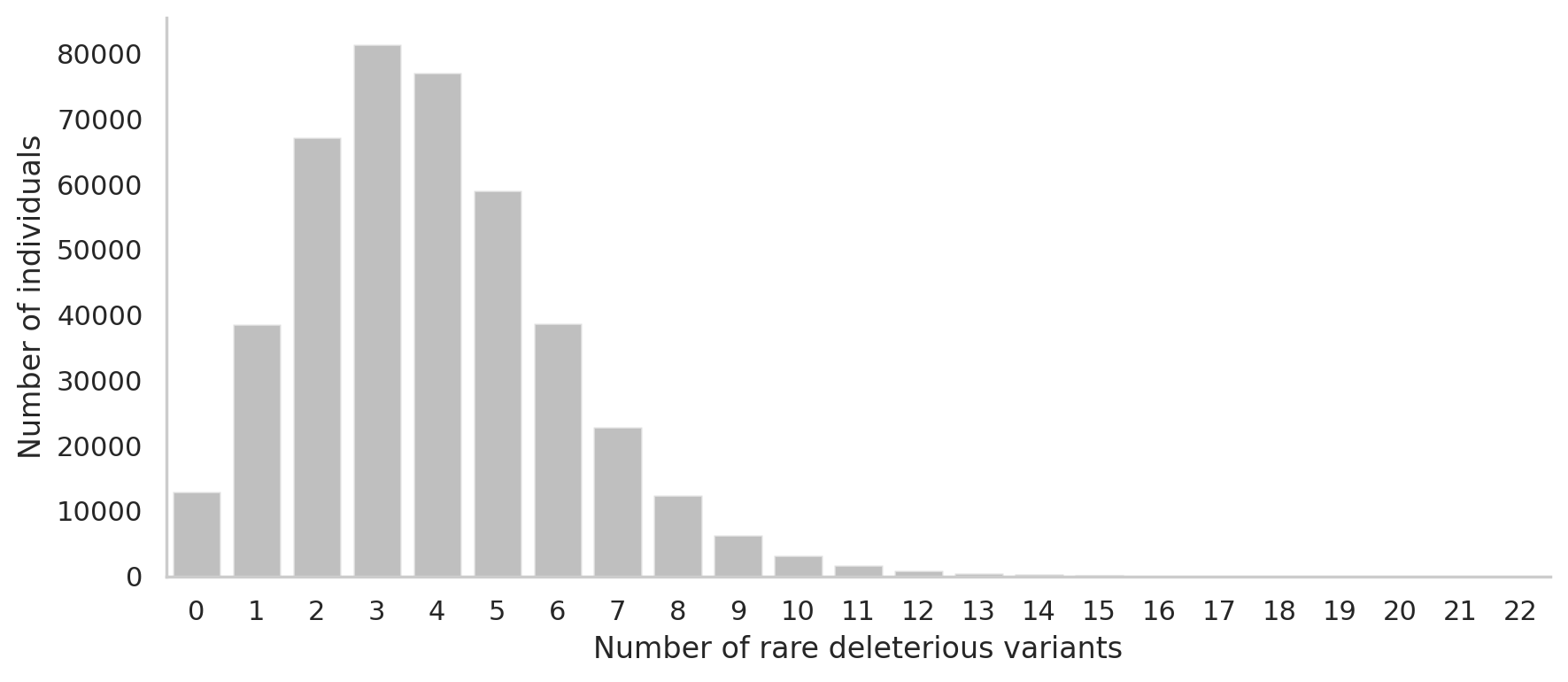

**Fig S13. Distribution of number of deleterious variants per individual across 90 traits.** Number of deleterious variants per individual in any of the genes from the gene-phenotype pairs that were significant at 5% FDR. There are 1,343 unique genes from 3,035 gene-phenotype pairs that passed the significance threshold. The average number of deleterious variants per person is 3.89 and the standard deviation is 2.21. 97% of all individuals have at least one deleterious variant.

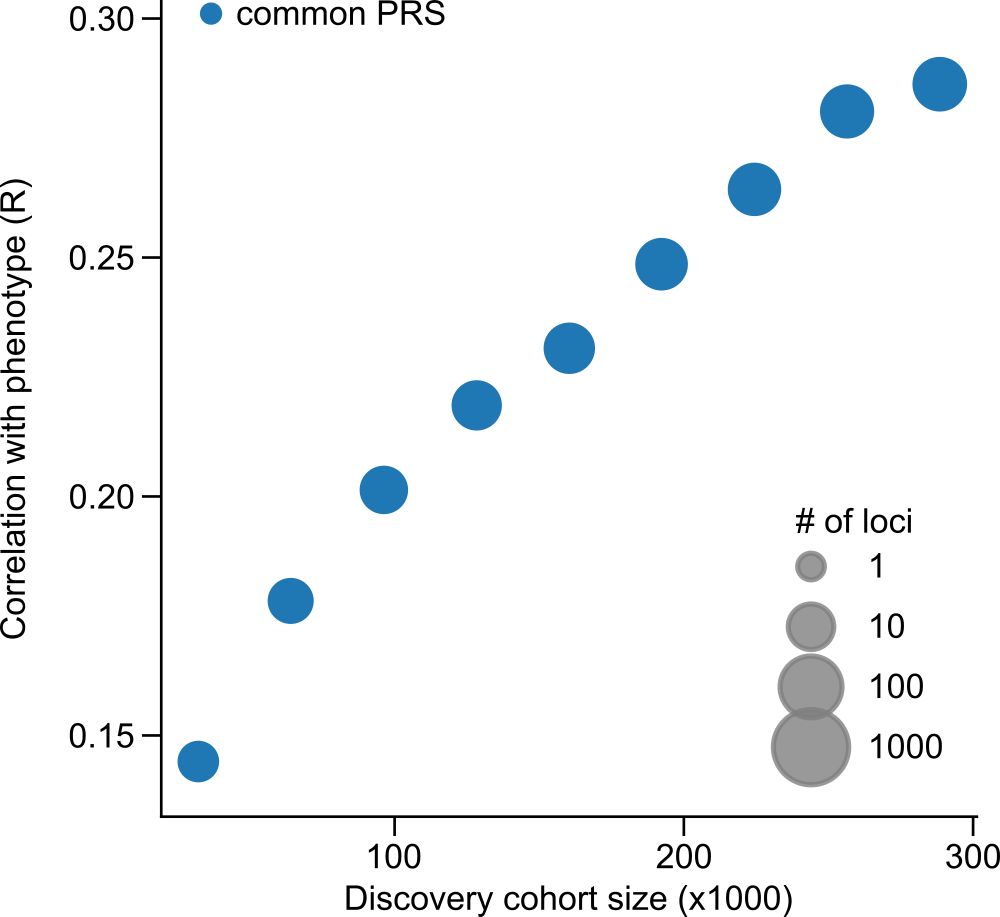

**Fig S14. Common PRS performance by training cohort size.** Improvement in common PRS performance with increasing discovery cohort size, measured by median correlation with phenotype.

**Fig. S15**. **Enrichments of rare variants in GWAS genes for all pairwise comparisons between 90 quantitative and clinical phenotypes**. For each pair of phenotypes, the heatmap shows the magnitude (size of circles) and statistical significance (color intensity) of the association of GWAS genes for phenotypes on the x-axis with rare variants affecting phenotypes on the y-axis. The burden tests for rare deleterious variants are computed after regressing out the effects of all significant GWAS variants from each phenotype. Fig. 4F in the main text shows a subset of the phenotypes which is enriched on the main diagonal in this figure.

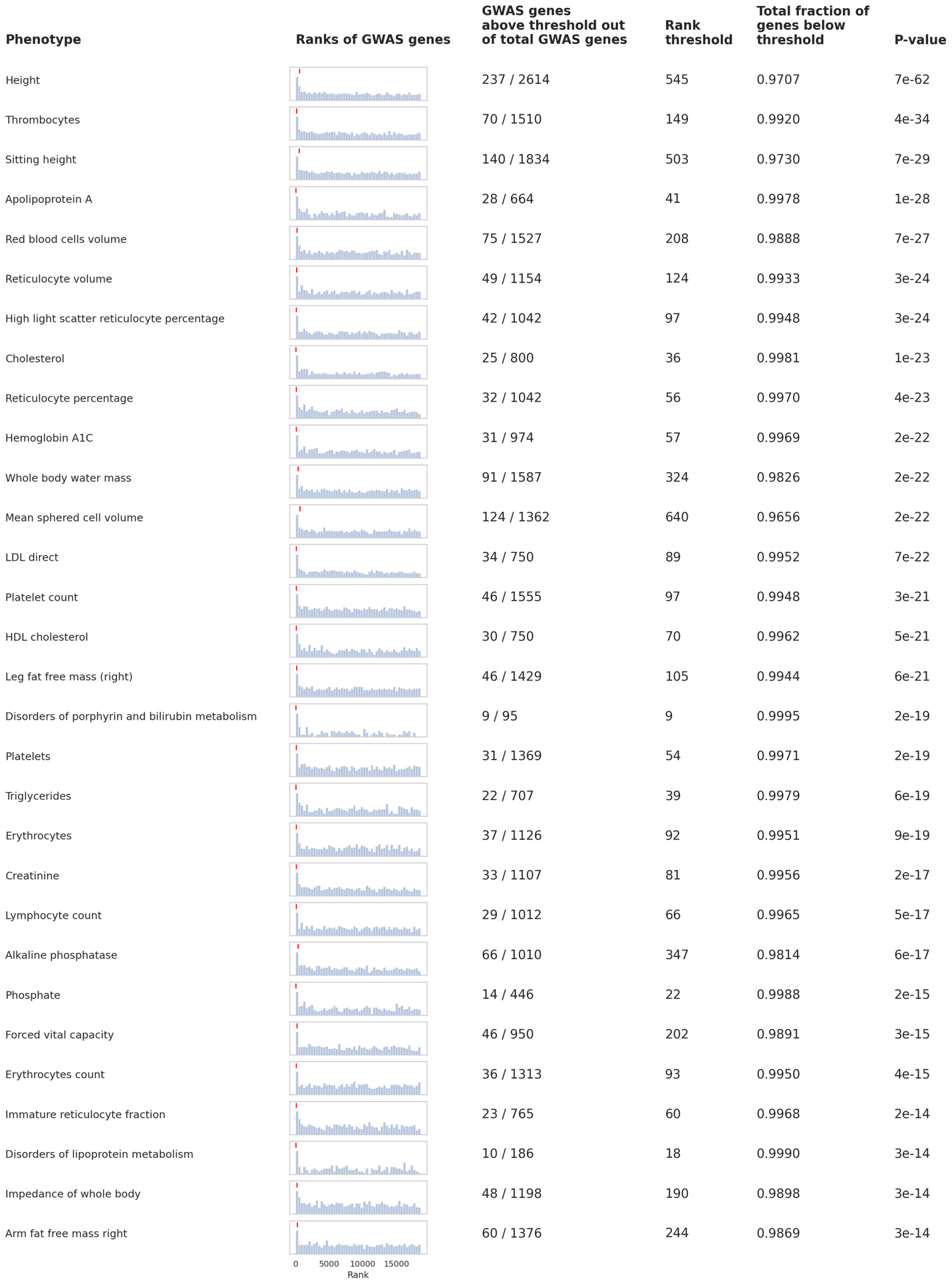

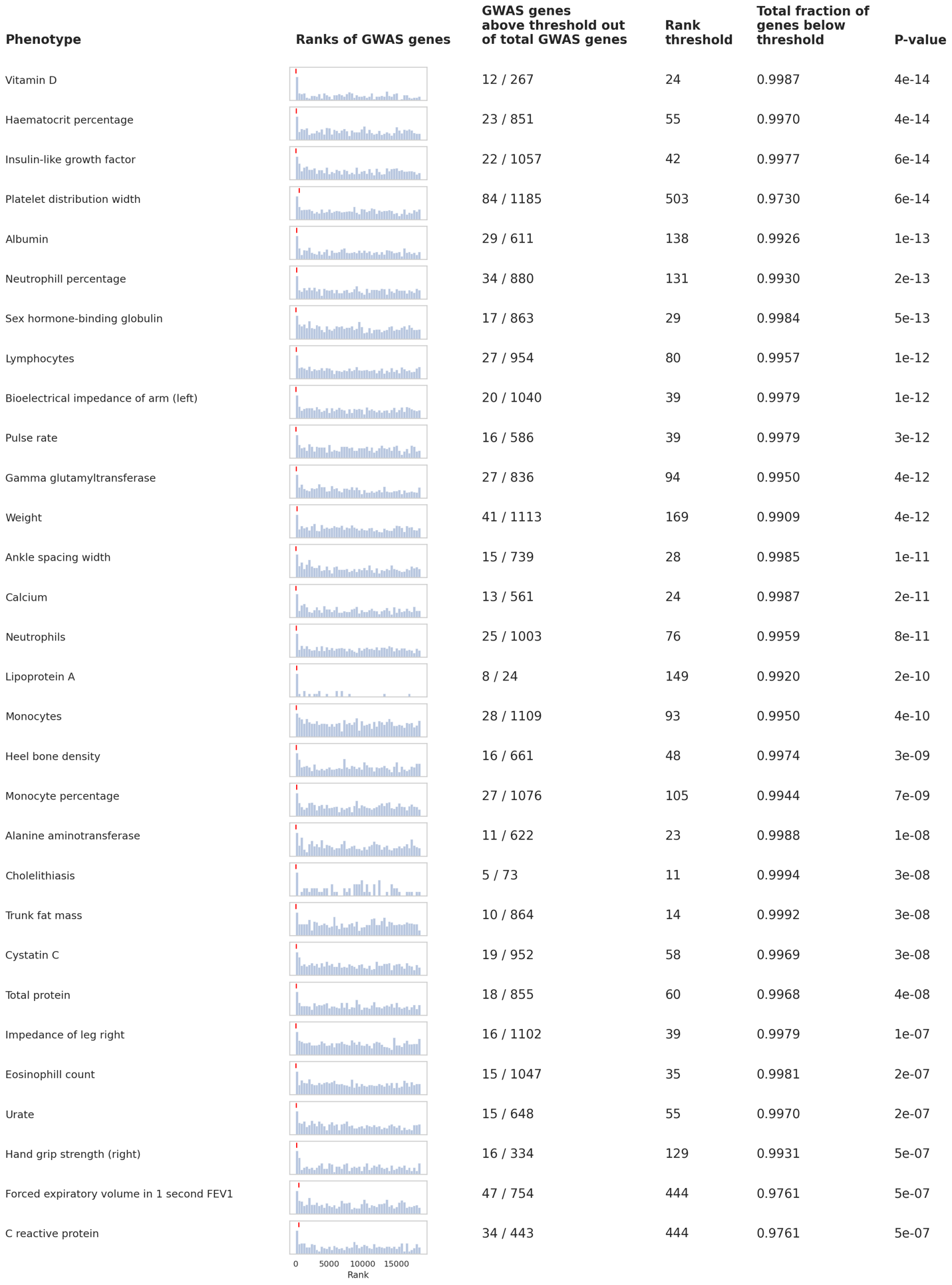

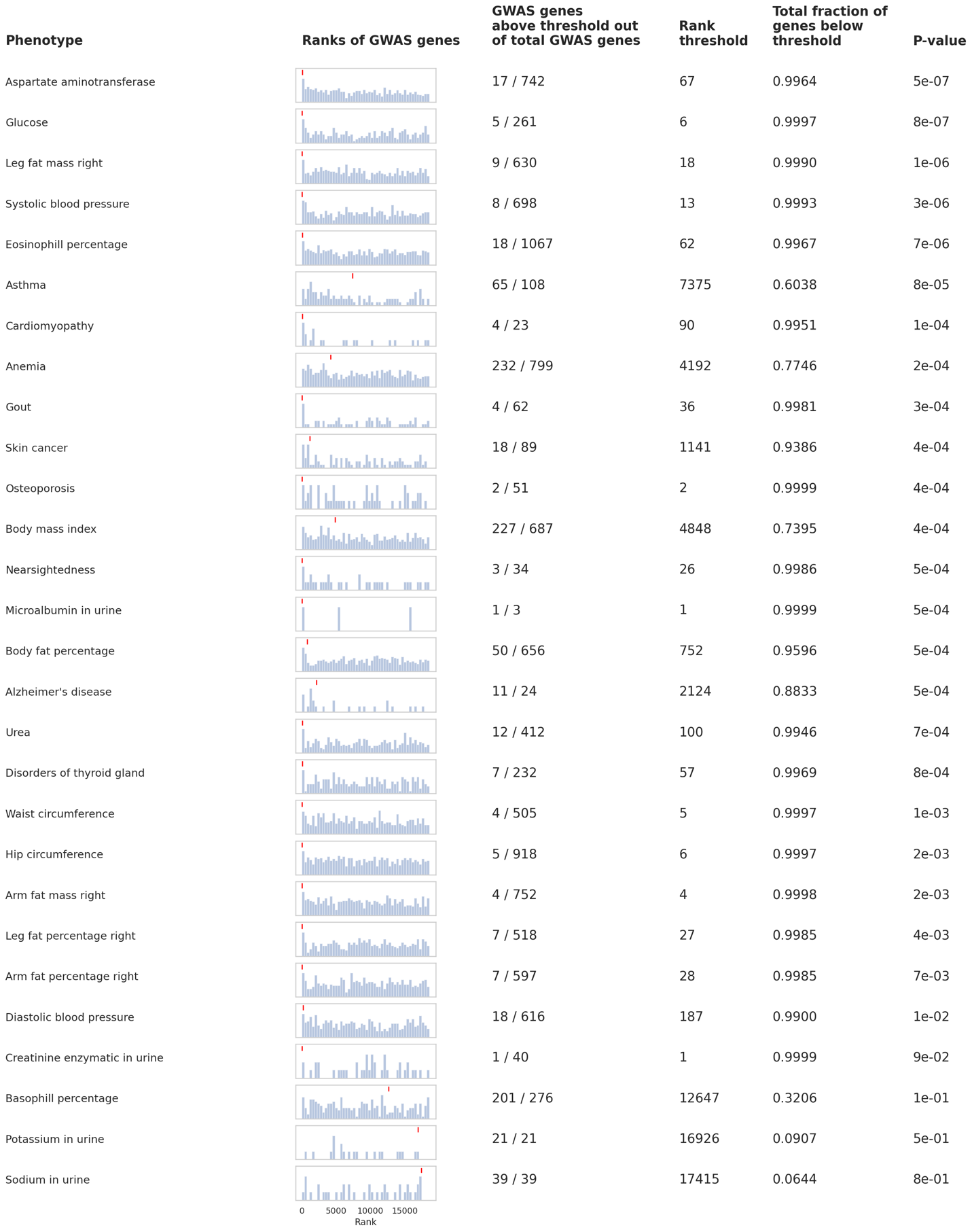

**Fig. S16. Subsets of GWAS genes for each phenotype that rank significantly high among all protein-coding genes based on the statistical significance of their enrichment for rare deleterious variants.** For each phenotype, a histogram of the ranks of all GWAS genes by their P-value from the rare variants burden test across all genes in the genome is shown. The red mark above the histogram indicates the rank threshold for the most significant subset.

### Supplementary table captions

**Supplemental Table S1**. Quantitative and clinical phenotypes from UK Biobank used in this study.

Columns: Phenotype: Name of analyzed trait; UKB field ID: UK Biobank data field ID corresponding to trait; # individuals in WES: Number of individuals measured for trait, and with whole exome sequence data available; # individuals in GWAS: Number of individuals measured for trait, and with array-based genotypes available. Several traits were insufficiently powered in the UK Biobank, and significant variants were instead obtained from the GWAS Catalog.

**Supplemental Table S2**. Categories of medications used to correct phenotypic values for drug use.

Columns: Category name: Broad category for medication type; Category ATC code: Anatomical Therapeutic Chemical (ATC) code for medication category; UKB medication name: Name of medication, as used in the UK Biobank; UKB medication coding: Medication code, as used in the UK Biobank; Medication ATC codes: ATC codes for medication; Common names: Commonly used names for medication; # UKB individuals on medication: Number of individuals in UK Biobank receiving the specific medication; # UKB individuals in category: Number of individuals in broader ATC medication category corresponding to the medication.

**Supplemental Table S3**. Covariates used to correct phenotypic values.

Columns: Covariate: Name of covariate; UKB field ID: UK Biobank field ID for covariate; Regressed from: Indicates which trait the covariate was regressed out from.

**Supplemental Table S4**. Gene-phenotype pairs which were significant on the rare variant burden test.

Columns: Phenotype: Named of analyzed trait from gene-phenotype pair; Gene: HGNC gene symbol for significant gene; Effect size: Effect size of carrying rare variants within the gene with respect to the normalized phenotype; P-value: P-value for gene-phenotype rare variant burden test; FDR: P-value, after correction for multiple testing with Benjamani-Hochberg false-discovery rate correction; UK Biobank AC threshold: Per-gene and per-phenotype allele count threshold used to define rare variants based on non-related individuals with phenotypic values in the UK Biobank; PrimateAI-3D percentile threshold: Per-gene and per-phenotype PrimateAI-3D threshold used to define deleterious missense; # carriers: Frequency of individuals carrying a rare deleterious variant in the gene, in the UK Biobank; # variants: Number of rare variants from gene used in burden test.

**Supplemental Table S5.** Independent GWAS variants at genome-wide significance for the 90 traits.

Columns: Trait: Name of trait; Chrom: Chromosome the variant is located on; Position (GRCh37): Nucleotide position of variant along the chromosome (for GRCh37 build); Reference allele: Reference allele for the variant; Alternate allele: Alternate allele for the variant; Effect size: Effect size for alternate allele of the variant with respect to the trait; P-value: P-value for association test between the variant alternate alleles and the phenotype; Allele frequency: Allele frequency of the alternate allele in the UK Biobank.

**Supplemental Table S6.** Number of significant genes identified by rare variant burden testing in this study and Backman *et al*. 2021.

Columns: Phenotype: Name of trait; # Genes in Fiziev, McRae et al.: Number of significant genes identified by rare variant burden testing by this study; # Genes in Backman et al.: Number of significant genes identified by rare variant burden testing by Backman *et al*. 2021.

**Supplemental Table S7.** Assessment of OMIM overlap in genes identified for UK Biobank phenotypes.

Columns: UKB Phenotype: Name of trait; OMIM Phenotype: Name of matched OMIM phenotype; MIM Number: OMIM catalogue number; Gene: HGNC symbol; Match: Indicator variable for phenotype match between UK Biobank and OMIM, 0=false, 1=true.

**Supplemental Table S8**. Gene-phenotype pairs used for evaluation of methods for variant prioritization.

Columns: Gene: HGNC gene symbol for gene; Phenotype: Name of significantly associated trait; Chromosome: Chromosome the gene is located on; LoF/p-value: P-value for rare variant burden test for LoF variants; LoF/beta: Effect size for rare loss-of-function (LoF) variants; LoF/n_carriers: Number of individuals carrying a rare LoF variant genotype in the gene, in the UK Biobank; LoF/n_variants: Number of rare LoF variants in the gene, in the UK Biobank; Missense/p-value: P-value for rare variant burden test for rare deleterious missense variants; Missense/beta: Effect size for rare deleterious missense variants; Missense/n_carriers: Number of individuals carrying a rare deleterious missense genotype in the gene, in the UK Biobank; Missense/n_variants: Number of rare deleterious missense variants in the gene, in the UK Biobank; Has >=10 ultra-rare Non-European missense variants: indicator variable, 0=False, 1=True; Has >=10 missense variants with at least 4 carriers: indicator variable, 0=False, 1=True.

**Supplemental Table S9**. Gene-phenotype pairs for which PrimateAI-3D scores of missense variants correlate significantly with phenotype values of their carriers.

Columns: Phenotype: Name of significantly associated trait; Gene: HGNC gene symbol for gene; Spearman R: Spearman correlation between PrimateAI-3D scores of rare missense variants and the average phenotype value across their carriers; # Carriers: Number of individuals carrying rare missense variants in the gene, in the UK Biobank; # Variants: Number of rare missense variants in the gene, in the UK Biobank; Nominal P-Value: P-value for the Spearman correlation, without multiple testing correction; FDR: P-value, after correction for multiple testing with Benjamani-Hochberg false-discovery rate correction.

**Supplemental Table S10**. Genes significant by rare variant burden test, used in construction of rare variant polygenic risk scores, by phenotype. Genes, effect sizes and P-values were obtained from analyses using individuals with exome data available (100% train).

Columns: Phenotype: Name of trait; Gene: HGNC gene symbol for the gene; Carrier Freq: Proportion of cohort carrying a rare deleterious variant given the AF and PrimateAI-3D thresholds; AF threshold: Alternate allele frequency threshold used to define rare variants per gene, and per trait; PrimateAI-3D threshold: PrimateAI-3D threshold used to define rare deleterious missense variants per gene, and per trait; Effect size: Effect size of carrying rare deleterious variants in the gene versus the trait; P-value: P-value from rare variant burden test between the gene and the trait;

**Supplemental Table S11**. Independent common variants, used in construction of common variant polygenic risk scores in UK Biobank, by phenotype. Variants, effect sizes and P-values were obtained from analyses using individuals with European ancestry and with exome data available (100% train).

Columns: Phenotype: Name of trait; Chrom: Chromosome the variant is located on; Position (GRCh37): Nucleotide position of the variant along the chromosome (for GRCh37 build); Minor: Minor allele for the variant; Major: Major allele for the variant; Effect size: Effect size of a minor allele against the trait; P-value: P-value from testing for association between minor allele dosage and the trait.

**Supplemental Table S12**. Characteristics of MGB cohort for rare variant PRS reproducibility.
